# A program for real-time surveillance of SARS-CoV-2 genetics

**DOI:** 10.1101/2024.04.18.24306026

**Authors:** Hayden N. Brochu, Kuncheng Song, Qimin Zhang, Qiandong Zeng, Adib Shafi, Matthew Robinson, Jake Humphrey, Bobbi Croy, Lydia Peavy, Minoli Perera, Scott Parker, John Pruitt, Jason Munroe, Rama Ghatti, Thomas J. Urban, Ayla B. Harris, David Alfego, Brian Norvell, Michael Levandoski, Brian Krueger, Jonathan D. Williams, Deborah Boles, Melinda B. Nye, Suzanne E. Dale, Michael Sapeta, Christos J. Petropoulos, Jonathan Meltzer, Marcia Eisenberg, Oren Cohen, Stanley Letovsky, Lakshmanan K. Iyer

**Affiliations:** Labcorp Center for Excellence in Data Science, AI and Bioinformatics, Burlington, NC 27215, USA; Labcorp Information Technology, Burlington, NC 27215, USA; Labcorp Research and Development, Burlington, NC 27215, USA; Labcorp Consumer Genetics Department, Burlington, NC 27215, USA; Labcorp-Sequenom, San Diego, CA 92121, USA; Labcorp Center for Esoteric Testing, Burlington, NC 27215, USA; Labcorp-Monogram Biosciences, South San Francisco, CA 94080, USA; Labcorp Drug Development, Burlington, NC 27215, USA

## Abstract

The COVID-19 pandemic brought forth an urgent need for widespread genomic surveillance for rapid detection and monitoring of emerging SARS-CoV-2 variants. It necessitated design, development, and deployment of a nationwide infrastructure designed for sequestration, consolidation, and characterization of patient samples that disseminates de-identified information to public authorities in tight turnaround times. Here, we describe our development of such an infrastructure, which sequenced 594,832 high coverage SARS-CoV-2 genomes from isolates we collected in the U.S. from March 13^th^ 2020 to July 3^rd^ 2023. Our sequencing protocol (‘Virseq’) generates mutation-resistant sequencing of the entire SARS-CoV-2 genome, capturing all major lineages. We also characterize 379 clinically relevant SARS-CoV-2 multi-strain co-infections and ensure robust detection of emerging lineages via simulation. The modular infrastructure, sequencing, and analysis capabilities we describe support the U.S. Centers for Disease Control national surveillance program and serve as a model for rapid response to emerging pandemics at a national scale.

## Introduction

The rapid emergence of COVID-19 and looming burden on global healthcare systems warranted swift responses from the international community. The causal virus, SARS-CoV-2, was first identified by metagenomic RNA sequencing^1^ as well as Sanger- and PCR-based detection methods^2,3^. Very early in the pandemic response, we prioritized the development of SARS-CoV-2 diagnostic assays to meet the demand for detection methods, offering one of the first PCR-based tests and performing as many as 275,000 tests daily^4^. This immense scale of PCR testing enabled us to assess the dynamics of COVID-19 infection as it pertains to PCR positivity^5^ and also provide population-based analysis on the maintenance of antibody titers^6,7^.

Similar to other betacoronaviruses, the SARS-CoV-2 genome mutated as it infected and spread across the population, with a mutation rate of approximately 1-2 x 10^-6^ mutations per nucleotide per replication cycle^8^. Such genetic changes are known to impact the severity and transmissibility of infection as well as vaccine efficacy^9^, thus requiring close to real-time surveillance using next generation sequencing (NGS)-based methods to inform public health policies. Multiple whole genome sequencing approaches have been applied to support this need, namely the ARTIC SARS-CoV-2 amplicon-based protocol for whole genome sequencing^10^, direct RNA sequencing^11^, and sequence hybridization^12^. Shotgun metagenomic sequencing of wastewater has also been an effective surveillance strategy for approximating variant abundances^13^ and identifying so-called ‘cryptic lineages’^14,15^, as it became apparent that COVID-19 patients exhibit fecal viral shedding^16,17^.

Ultimately, a greater need for high-throughput, real-time genomic sequencing of SARS-CoV-2 emerged in the United States through the SARS-CoV-2 Sequencing for Public Health Emergency Response, Epidemiology and Surveillance (SPHERES) spearheaded by the Centers for Disease Control (CDC). To address this public health need and in collaboration with CDC SPHERES, we rapidly developed a national surveillance apparatus using our purposely designed infrastructure for flexible sampling, a unique approach for SARS-CoV-2 whole genome sequencing, and tailored analytical methodologies that ensure continuously robust SARS-CoV-2 lineage determination using the Phylogenetic Assignment of Named Global Outbreak (PANGO) nomenclature^18^. Our assay, which we call ‘Virseq’, is distinguished from other NGS-based SARS-CoV-2 whole genome sequencing approaches through its use of probe-based tiling^19^ and long reads, which provide versatile, mutation-resistant capabilities. As of July 3^rd^, 2023, we have sequenced 594,832 genomes (10X median depth of coverage) and have provided 524,498 high-quality SARS-CoV-2 genomes (10X median depth of coverage, >90% genome coverage, complete S gene coverage) and patient demographic data to the CDC using our Virseq assay, representing continuous snapshots of SARS-CoV-2 viral evolution.

In this study, we provide a retrospective analysis of our Virseq assay using our vast repertoire of high-quality genomes, showcasing our surveillance capabilities and the modular resources that support its continued use. We show that Virseq has generated uninterrupted surveillance reflecting the nationwide prevalence of SARS-CoV-2 consistent with our RT-PCR sample collection, and we further demonstrate the robustness of SARS-CoV-2 lineage determination through our in-house analytical capabilities. We also address the analytical challenges posed by rare SARS-CoV-2 co-infections by developing a custom workflow that yields haplotype-resolved consensus genomes. Finally, we pose this suite of resources as a model for mounting rapid and robust large-scale surveillance networks.

## Results

### An infrastructure for nationwide COVID-19 surveillance

The rapid emergence and continuous evolution of SARS-CoV-2 necessitated setting up a national surveillance program that could provide real-time epidemiological snapshots across the United States. In response to the CDC’s basal surveillance program, we organized and implemented a nationwide infrastructure to identify SARS-CoV-2 positive patient samples across the United States, consolidate these samples and their associated demographic data, and sequence their genomes to determine their SARS-CoV-2 PANGO lineages. Our surveillance system also includes mechanisms for monitoring emerging lineages and their potential impacts on qPCR and sequencing performance. Further, this setup is flexible and modular, enabling rapid responses and developments to our pathogen surveillance.

A schematic of this modular infrastructure and its associated sequencing protocols is shown in **Figure 1**. In **Figure 1a**, we show a high-level view of our surveillance pipeline that begins with sample accessioning and resulting in whole genome sequencing reports with de-identified sample metadata. Upper respiratory samples (nasopharyngeal (NP) or nasal swabs) collected in 0.9% saline or viral transport media were collected and analyzed through our COVID-19 RT-PCR Test at various Labcorp^®^ service centers and laboratories, and the resulting PCR extraction plates containing SARS-CoV-2-positive samples were then shipped to Labcorp^®^ central locations and consolidated into plates with high viral titer samples (i.e. N1 Ct < 31) using our custom developed plate selector app (**Figure 1b**, Methods). The selection of plates using this app is critical, as it de-identifies data thereby ensuring HIPPA compliance and allows flexibility in choosing samples. For example, we can focus on certain geographic regions where outbreaks are underway, put limitations on the amount of data from a state to ensure uniform surveillance, and threshold on the PCR amplification N1 Ct value to ensure sufficient genetic material is available for sequencing in each sample. The resulting consolidated plates of high viral titer samples were then shipped to our sequencing labs where samples were processed through custom tiled Molecular Loop^®^ probe amplification, followed by library preparation and sequencing on the PacBio^®^ Sequel II™ platform in a highly multiplexed fashion (**Figure 1c-d**). PacBio^®^ raw data was then processed to generate Circular Consensus Sequencing (CCS) reads which were then analyzed using our custom bioinformatics workflow to generate consensus genomes for each sample (**Figure 1d**). Stringent sequence coverage quality control was then applied followed by PANGO lineage determination for each sample (Methods), and results were then merged with patient demographic data and de-identified with a new custom ID generated for each sample (**Figure 1d**). Consensus genomes, summary reports, raw CCS reads, and alignment variant calls were then provided to CDC who in turn processed our submission and deposited the relevant data to public repositories, namely GISAID^20^ and NCBI^21^.

**Figure 1.**
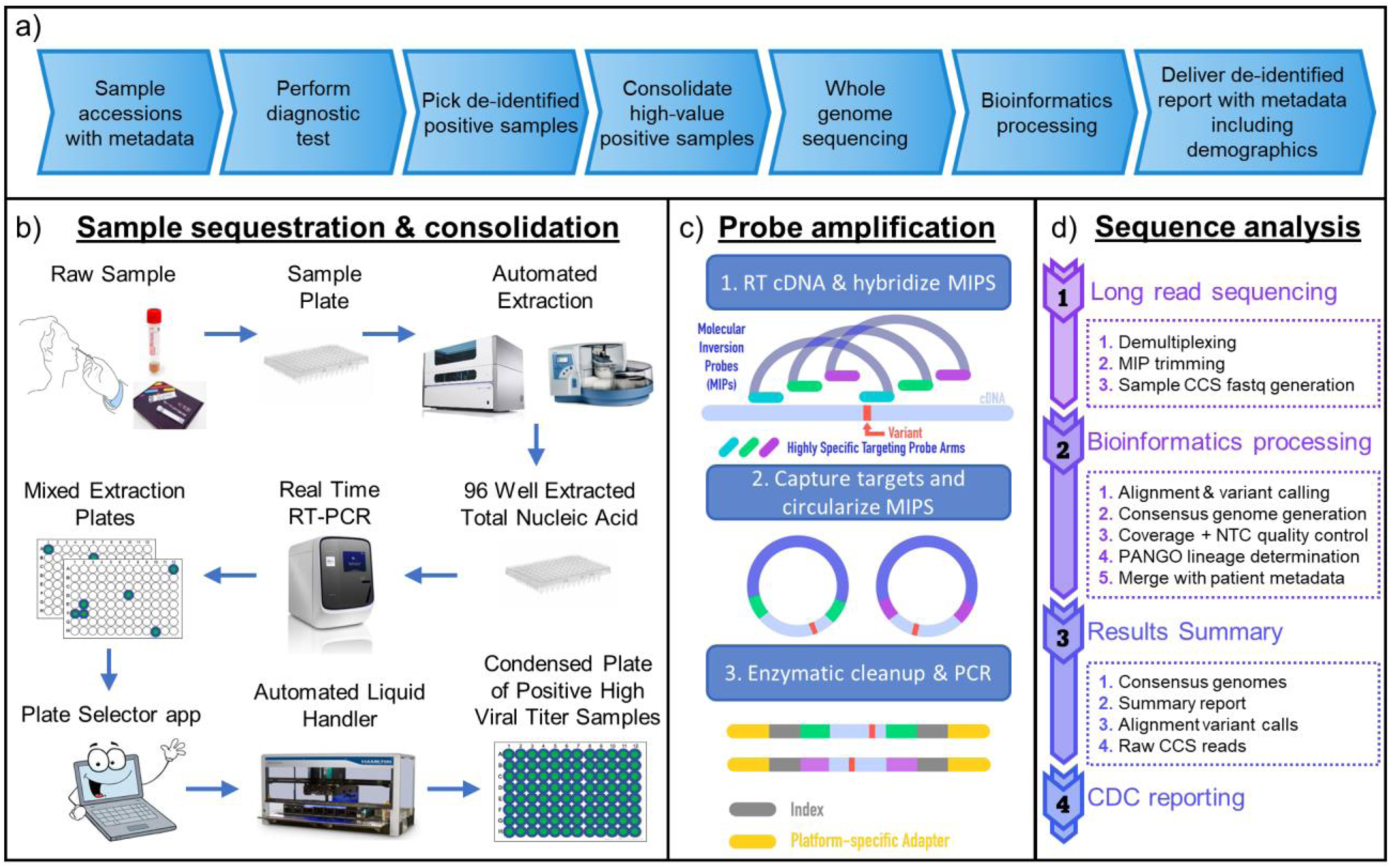
Journey of a sample from raw nasopharyngeal and nasal swabs to CDC reporting. a) Overview of end-to-end genomic surveillance setup. b) Consolidation of positive high viral titer samples. c) Probe amplification protocol. d) Long read sequencing analysis and bioinformatics workflow to prepare high-quality SARS-CoV-2 genomic sequences with demographic metadata for CDC.

### High-throughput, high-fidelity SARS-CoV-2 genome surveillance pipeline

The rapid pathogen evolution during a pandemic and the possibility of sporadic outbreaks necessitates a highly robust genomic surveillance pipeline. From January 2021 to July 2023, we have used our Virseq pipeline (**Figure 1**) to report 524,498 high-quality SARS-CoV-2 genomes (10X median depth of coverage, >90% genome coverage, complete S gene coverage) and patient demographic data to the CDC. These sequences captured all major lineages that have emerged throughout the COVID-19 pandemic since the inception of this surveillance effort, including Alpha, Delta, Omicron, and the many Omicron subvariants (**Figure 2**, Methods). When overlaying the positivity rate of our diagnostic PCR assays used for sample picking, we observed multiple fluctuations matching variant emerges, such as BA.1, BA.4/BA.5, and XBB.1.5 (**Figure 2b**). Stratifying genomes by U.S. Health and Human Services (HHS) region (HHS regions 1-10; Methods), we found that the HHS regions 1 and 2 (corresponding to the U.S. Northeast) often served as a harbinger to predict variant emergence for all other regions (**Figure S1**). For example, the initial Omicron variant (BA.1) and the more recent XBB.1.5 variant reached ∼50% prevalence in HHS regions 1 and 2 approximately one week and four weeks prior to all other regions, respectively (**Figures S1 and S2**).

**Figure 2.**
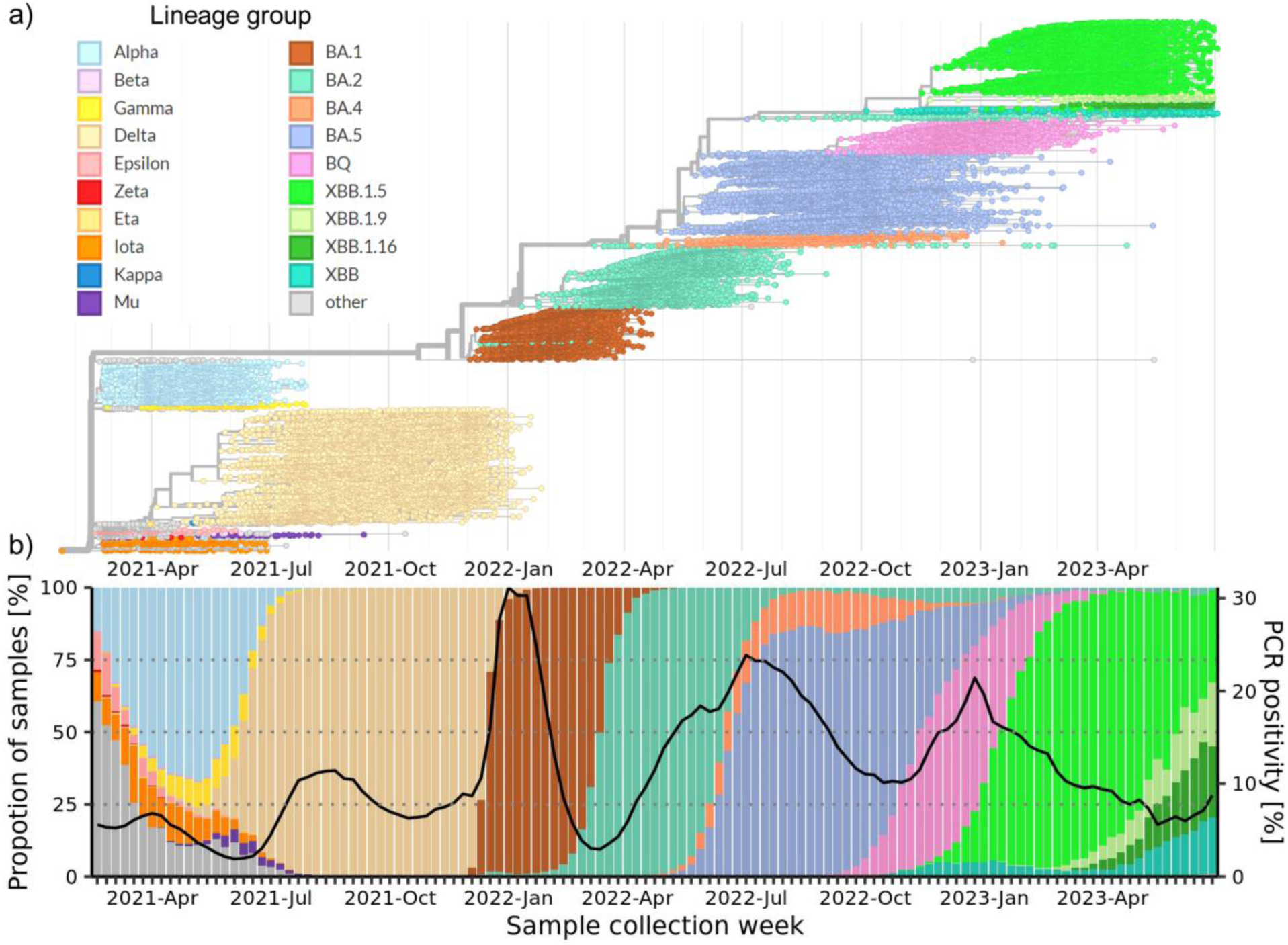
SARS-CoV-2 PANGO lineage analysis of 594,832 high-quality genomes from USA samples collected between January 2021 and July 2023. In each plot samples are grouped and colored by the closest parent listed on the top left. a) Time-resolved phylogeny of a subset of samples (n=28,069) clustered based on the oldest collected sample with at most 1,000 samples per month. b) SARS-CoV-2 lineage proportions across weeks with at least 100 samples (left y-axis) and the PCR positivity rate (%) indicated by the black line (right y-axis).

After the BA.1 wave, reports indicated that SARS-CoV-2 infections by Omicron variants exhibited lower viral loads^22,23^, prompting us to investigate the diagnostic PCR N1 Ct values of our samples, which are a useful proxy for viral load. As expected, lower sample Ct values were correlated with both increased average depth of coverage and higher consensus genome coverage (**Figure S3a**). We also observed shifts in sample diagnostic N1 Ct values throughout the pandemic that ranged from 20-21 in 2021 and exceeded 24 during the BA.1 wave in the 2021-2022 winter season, prior to reaching a steady state between 22-23. (**Figure S3b**). In our later analysis of SARS-CoV-2 co-infections, we found that co-infected sample N1 Ct values followed the same trends as those of samples where only a single SARS-CoV-2 lineage was detected. Together, these results indicate that our surveillance captures the overall kinetics and patterns of circulating variants.

This collection of high-quality genomes also uniquely captured demographic trends from all 50 states in the U.S. and the District of Columbia. States with the highest representation include California (n>62,000) and New Jersey (n>48,000), while other key states from HHS regions 9 and 10 (WA, n >19,000; AZ, n>17,000), HHS region 5 (IL, n>23,000; OH, n>10,000), and HHS regions 4 and 6 (NC, n>45,000; FL, n>36,000; TX, n>14,000) also had strong sampling (**Figure 3a**, **Table S1**). We also observed that the number of Virseq-generated genomes has represented a consistent proportion of total SARS-CoV-2-positive samples collected in each HHS region with minor fluctuations (**Figure 3b**). These fluctuations are expected as the timing of variant outbreaks (e.g. BA.1) can vary across HHS regions along with concomitant surges in PCR positivity rate (**Figure 2b**). Despite these changes, we still observed sustained census normalized sampling across both our diagnostic PCR assays and Virseq (**Figure S4a**) even with an end to the COVID-19 emergency response and the reduction in our surveillance volume in 2023 (**Figure S4b**). Notably, we observed a reduction in Virseq surveillance in HHS regions 6-8 at the onset of the BA.2 wave, as BA.2 prevalence spiked elsewhere in the U.S. before affecting these regions (February 2022; **Figures 2b, 3b**, **S1, S2, and S4**).

**Figure 3.**
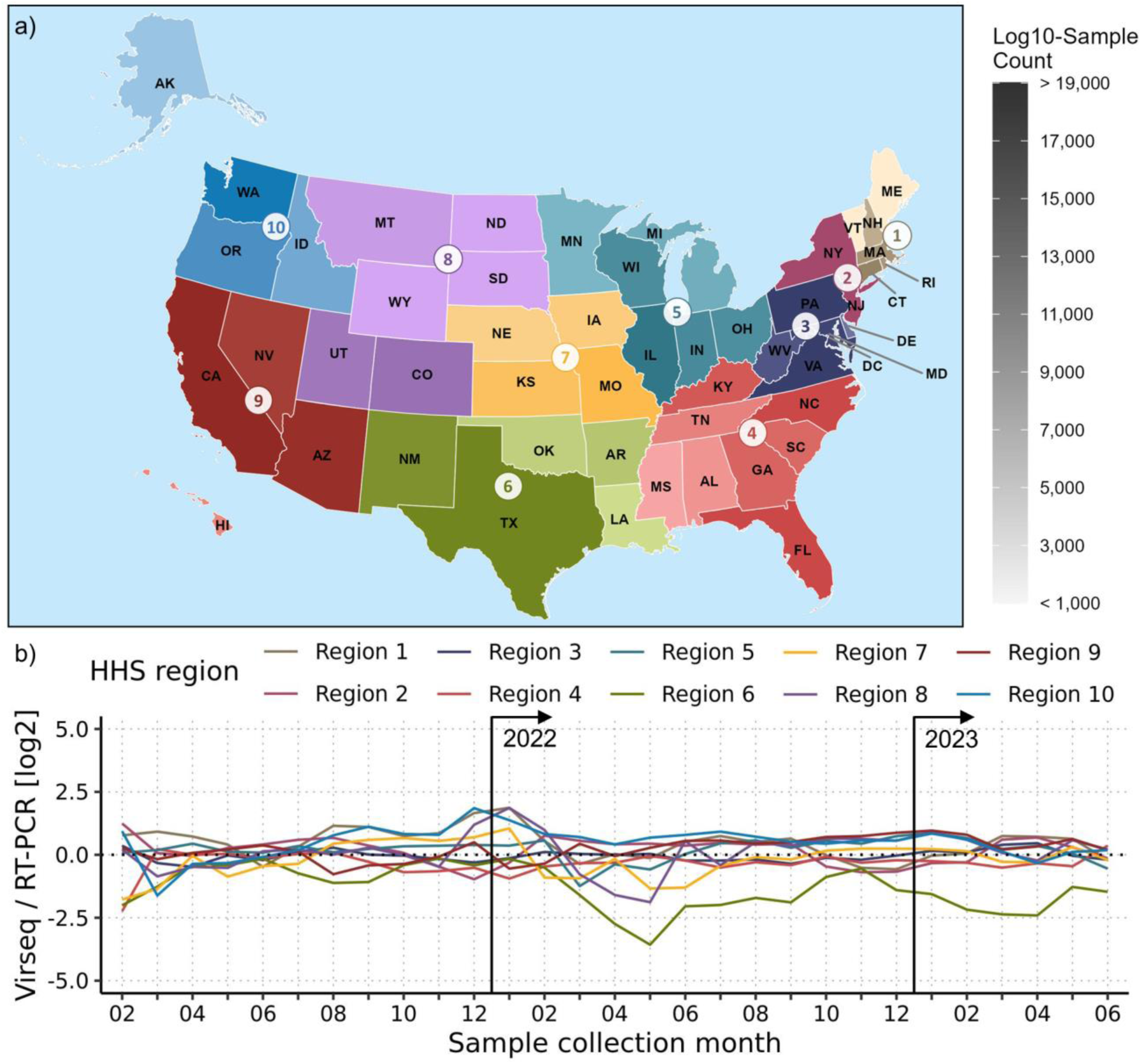
U.S. nationwide SARS-CoV-2 genomic surveillance consisting of 594,832 high-quality genomes from samples collected between January. 2021 **and July** 2023. In both panels, the U.S. is divided into 10 Health and Human Services (HHS) regions, each in separate colors. **a)** Each state is notated by its two-letter abbreviation and the shade indicates the number of samples collected. Each region uses a shared color gradient, shown on the right in grayscale with maximum and minimum values of 19,000 and 1,000, respectively. **b)** Line plots showing the ratio of Virseq-generated genomes versus the total SARS-CoV-2-positive RT-PCR samples collected on the log2 scale over each sample collection month. January 2021 and July 2023 were excluded, as they each have fewer than 100 Virseq-generated genomes in the dataset analyzed. Vertical black lines denote the start of years 2022 and 2023.

We also found that the age distribution of samples collected in each geographic region shifted throughout the course of the pandemic, with a plurality from pediatrics in 2021, shifting to a more even distribution of ages in 2022, and finally shifting to a plurality from older segments of the population in 2023 (**Figures S5 and S6**). Interestingly, there was also a modest trend in patient-reported gender, as the proportion of samples from female patients appeared to increase from 2021 to 2022 and once again in 2023 (**Figures S5 and S6**), despite there not being any difference in PCR positivity between males and females (**Figure S7a**). We also observed a bifurcation in PCR positivity across age groups in March 2022, as pediatric PCR positivity lowered to approximately half of the positivity in most other age groups and this relative difference has not changed since (**Figure S7b**). Intriguingly, two months prior (January 2022), the CDC and FDA announced multiple expansions of pediatric COVID-19 vaccination availability^24–26^ (**Figure S7b**). We also observed that in June 2022, 18–19-year-olds had a reduction in PCR positivity relative to older age groups as well, and most recently in June 2023 we observed an increase in geriatric (80+ year olds) PCR positivity (**Figure S7b**). These demographic- and region-specific trends in variant prevalences and PCR positivity rates require a surveillance apparatus like ours that is flexible and robust to the rapid evolution of the SARS-CoV-2 genome.

### Robust, mutation-resistant S gene sequencing using a probe-based long-read strategy

To maintain nationwide surveillance of pathogen genome evolution, the selected whole-genome sequencing approach must be able to withstand sudden changes in genetic diversity. Our surveillance apparatus uniquely employs a probe-based long-read sequencing approach that is mutation-resistant by design due to its ∼22X genome tiling of >99% of the SARS-CoV-2 genome, except for a few hundred base pairs of the 5’- and 3’ peripheral genomic regions (**Figure 1c**). At the end of 2021, the Omicron variant (BA.1) emerged and swept through the U.S. in a matter of weeks (**Figures 2**, **S1, and S2**), dramatically shifting the diversity of the circulating lineages across the U.S. population (**Figure 4a**). Unlike earlier variants like Delta which were predominantly mutated in the ORF1a genic region, the original Omicron variant (BA.1) introduced a surge of novel S gene mutations (27 SNPs and three deletions compared to Delta) (**Figure 4b**), raising concern regarding the ability of PCR- and amplicon-based assays to detect BA.1. In fact, the Spike gene target failure (SGTF) genomic signature was so common that it became a useful proxy for the PCR detection of emerging variants, such as Alpha and Omicron^27^. While interruptions in our surveillance were not observed (**Figure 2**), we verified the fidelity of our sequencing of BA.1* using an *in silico* approach to check for probe dropout caused by lineage defining mutations (Methods). We found that our assay retained ∼20X *in-silico* tiling of the S gene during the initial Omicron wave (**Figure 4c**), and in the worst-case scenario where we allowed zero SNP tolerance in probe binding regions, we retained a minimum of ∼10X probe tiling (**Figure S8**). Furthermore, as the total number of unique mutations and the concurrent prevalence of multiple circulating lineages measured in terms of their entropy continues to increase with more recent XBB subvariants, we continue to observe profoundly stable genome-wide probe tiling *in silico* (**Figure 4c**). This robust probe tiling is especially important as chronic infections and widespread vaccination have altered the evolutionary trajectory of the SARS-CoV-2 genome^28^ and the receptor binding domain of the S gene remains under considerable selective pressure in the Omicron era^8^. We also confirmed the robustness of our sequencing strategy by analyzing the genome-wide per-base coverage of Variants of Concern (VOCs) that have emerged throughout the pandemic (**Figure S9**, starting with B.1.1.7 or ‘Alpha’ up until XBB.1.5). We found that overall per-base coverage has remained stable and well above our minimum per-base coverage required for base calling in our consensus genomes, despite the heavily mutated S gene of Omicron and subvariants thereof (**Figure S9**). Together, these results show that the Virseq assay is a stable and effective sequencing strategy and is a critical component of our surveillance apparatus. However, while we are confident in our response to variant outbreaks thus far, it is imperative that proactive measures are taken to preclude future surveillance interruptions.

**Figure 4.**
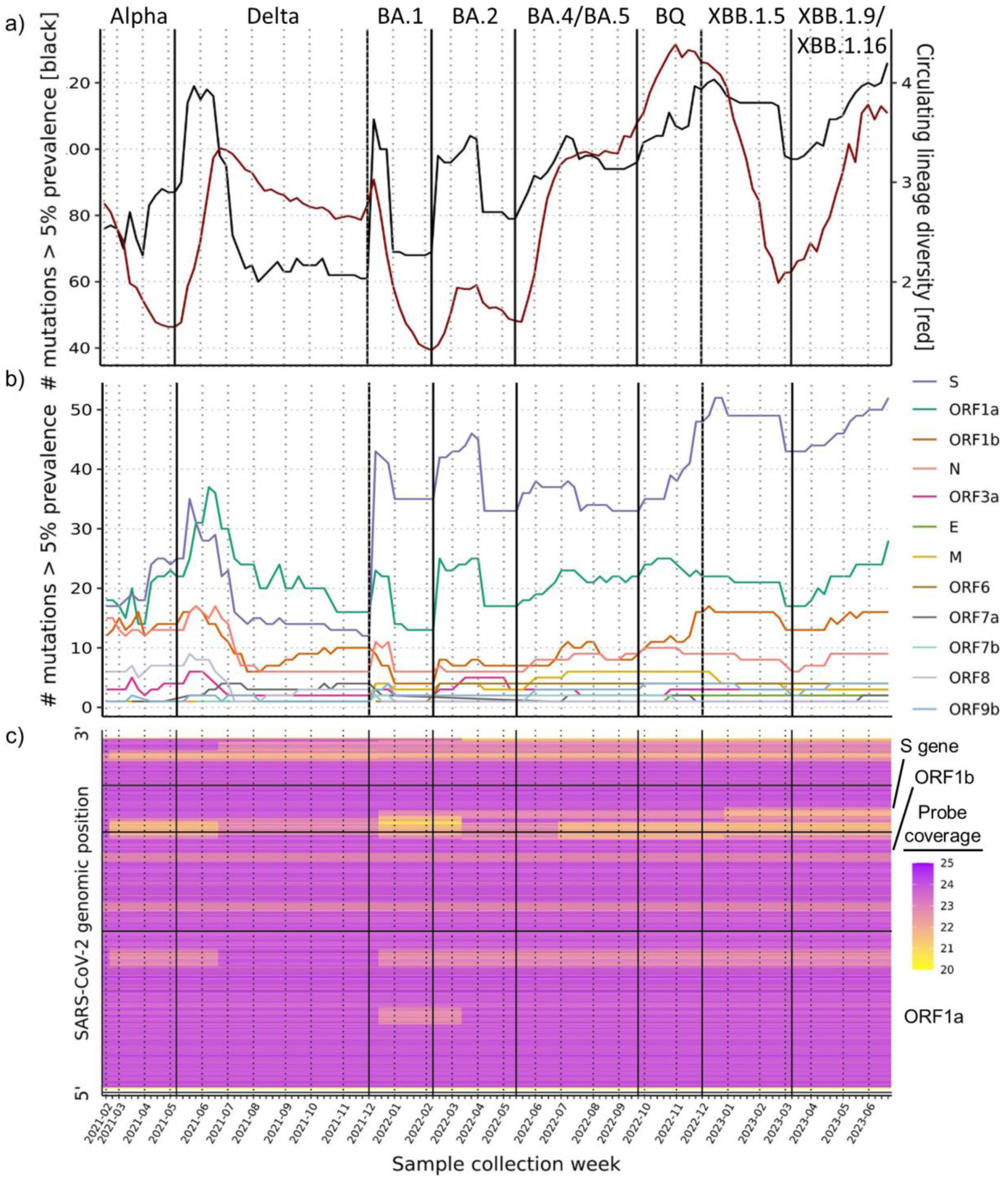
Pandemic-wide trends in SARS-CoV-2 genomic mutations and the robustness of whole genome probe-based sequencing. a) Number of mutations with at least 5% prevalence in the lineage population (black) and circulating lineage shannon diversity (red). **b)** Number of mutations with at least 5% prevalence in the lineage population separated by genic origin. **c)** Heatmap showing the genome-wide probe coverage of the most common lineage in circulation for each collection week with genomic positions shown 5’ (bottom) to 3’ (top). Probes were considered failures if a deletion, insertion, or >3 SNPs were detected in either probe arm. Large genic regions (ORF1a, ORF1b, S) are indicated by horizontal lines and are labeled on the right. In all panels, results are stratified by sample collection week with vertical bars separating the major waves of the pandemic with the causal variant shown above. Waves are demarcated using the collection week when the causal variant first reached 5% prevalence.

### Modeling the performance of the Virseq assay by simulation

One challenge of sustaining continuous whole genome surveillance is the need to predict changes in sequencing performance that may occur and its effect on the characterization of the pathogen variants. Many SARS-CoV-2 lineages emerge in regions outside of our surveillance network (i.e. in countries other than U.S.) and may be too rare for detection once they initially spread to our surveilled regions. To address this challenge, we developed a Virseq performance simulator that models our entire production process from raw reads to PANGO lineage determination, capturing the sequencing and other systematic errors that might propagate into consensus genomes. This simulator was constructed using a representative batch of samples and incorporates the per-base coverage and minor allele fractions commonly observed at each genomic position (Methods, **Figure S10**). Application of this simulator begins with an input sequence that is then mutated to reflect any errors introduced by our production process, which can then be compared with the original sequence via concordance analysis of their PANGO lineage determinations.

We routinely use this simulator to monitor newly designated lineages in the PANGO nomenclature (largely VOCs) as well as randomly selected sequences from previous months of surveillance, leveraging the GISAID database^20^. This routine monitoring mechanism is a crucial component of our FDA EUA and could be used to help maintain future pandemic surveillance networks. In this study, we expanded this analysis to include up to 100 sequences each from 1,899 VOCs (97,421 total sequences, ‘VOC experiment’) and descendant lineages thereof, current and former, and 10,000 randomly selected sequences from each month spanning from January 2021 until July 2023 (310,000 total sequences, ‘retrospective experiment’) (Methods). As expected, we observed similar coverage profiles between the two experiments and the simulator model, indicating that the simulated genomes accurately reflected Virseq-generated sequences (**Figure S10b**). When assessing the PANGO lineage concordance between the simulated and original genomes, we first checked for exact matches then also checked for parent/child relationships between the lineages compared (e.g. BA.5 is a parent of the child BA.5.1), deeming these parent matches (Methods).

Overall, we observed strong concordance in both the retrospective (99.55% exact, 99.97% parent) and VOC experiments (99.06% exact, 99.84% parent) (**Figure 5**). In the retrospective experiment we observed strong concordance across all 31 months analyzed with some month-to-month fluctuations (>98.95% exact, >99.9% parent) (**Figure 5a**). We also observed that some fluctuations coincided with shifts in circulating lineage diversity and the timing of VOC emergences (**Figure 4**, **Figure 5a**). Intriguingly, while some VOC emergences resulted in slight reductions of concordance (BA.1, BA.4/BA.5), others counterintuitively coincided with improved concordance (BA.2, BQ, XBB.1.9/XBB.1.16) (**Figure 4**, **Figure 5a**).

**Figure 5.**
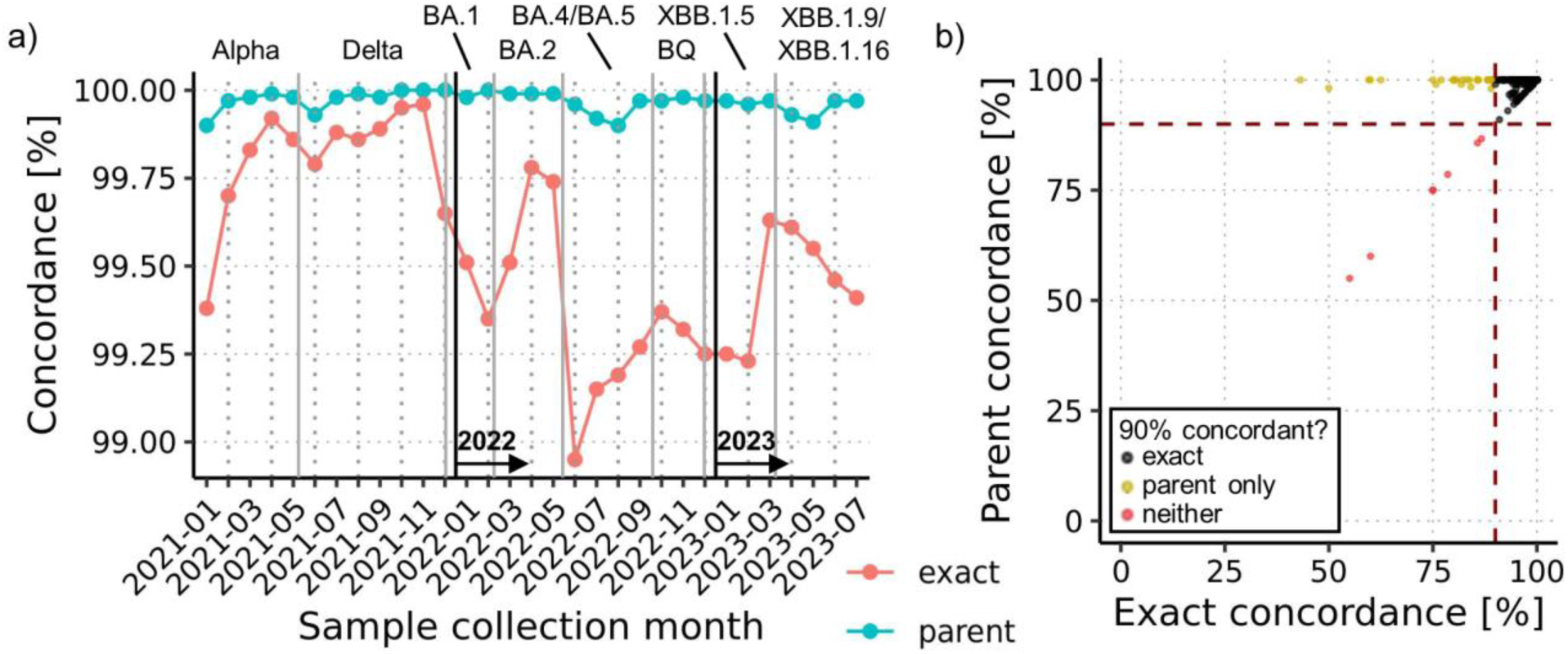
Virseq simulation results. Exact and parent concordance for the retrospective (**a**) and VOC (**b**) simulation experiments. Retrospective results are shown with exact (red) and parent (blue) concordance as separate lines over the sample collection months analyzed, with years demarcated by vertical lines. VOC results are shown with exact and parent concordance results for each VOC data point with 90% thresholds marked by dashed red lines. VOCs colored black have >90% exact concordance, those colored yellow only have >90% parent concordance, and those colored red are below both concordance thresholds. Vertical gray bars separate the major waves of the pandemic with the causal variant shown above. Waves are demarcated using the collection week when the causal variant first reached 5% prevalence.

When assessing the concordance of individual VOCs, we found that 98.21% (1,865) and 99.63% (1,892) of VOCs had >90% exact and parent lineage concordance, respectively (**Figure 5b**). In total, we observed seven VOCs with a small number of discordant calls, and four of these (BA.2.2.1, BA.5.10, BQ.1.19, and BY.1) had UShER tree placement conflicts with their designated hashes, i.e. these sequences were representative of those lineages but could not be placed accordingly in the UShER tree (**Table 1**). This indicated that the original sequences of these VOCs had unstable PANGO lineage designations. For example, one of four simulated BA.2.2.1 sequences was called a BA.1, and we later found that the original sequence was hashed as BA.2.2.1 but placed in the UShER tree as BA.1 (**Table 1**). Inspection of the other three VOCs revealed that they were recombinants with discordant calls corresponding to one of the recombined lineages (**Table 1**), indicating that the simulated genomes had a loss of resolution.

**Table 1.** Summary of VOCs with more frequent discordant PANGO lineage calls. Each row shows a lineage with < 90% parent concordance and the most common discordant lineage observed among the simulated sequences.

| PANGO lineage | Concordance (%) | Total sequences analyzed | Most common discordant lineage |
| --- | --- | --- | --- |
| **BA.2.2.1** | 75.00% | 4 | BA.1 |
| **BA.5.10** | 60.00% | 10 | BA.5.2 |
| **BQ.1.19** | 75.00% | 4 | BQ.1.2 |
| **BY.1\*** | 86.67% | 15 | BA.2.75.7 |
| **XB\*** | 55.00% | 100 | B.1 |
| **XBV\*** | 78.57% | 14 | XBB.1 |
| **XP\*** | 85.71% | 7 | BA.1.1 |

\* BY is the alias of BA.2.75.6. XB, XBV, and XP are the following recombinants, respectively: B.1.634/B.1.631, CR.1/XBB.1, and BA.1.1/BA.2.

We then investigated features of the simulated genomes, including key drivers of discordant lineage calls. As expected, we found that genome coverage was significantly lower among genomes that had discordant lineage calls or that were only parent concordant compared to those with exact concordance in both experiments (p<0.001 in all comparisons, Wilcoxon rank-sum test, **Figure S11a-b**). Both experiments yielded similar genomic positional dependence of consensus genome errors (**Figure S11c**), and these errors were often found in regions modeled with poor coverage (5’/3’ peripheral genomic regions) instead of positions modeled with higher base calling errors (**Figure S11d-e**). These simulation experiments collectively show that the Virseq assay generates consensus genomes with accurate PANGO lineage designations and that accuracy is predominantly driven by genomic coverage, which is expected behavior for the pangolin software in general^29^. This Virseq simulator is a crucial component of our surveillance apparatus, ensuring that we anticipate potential sequencing interruptions and serving as a model for viral sequencing simulators in general.

### Detection and haplotype phasing of SARS-CoV-2 mixtures

The concurrent circulation of multiple lineages of the same virus during a pandemic may result in co-infections of different viral lineages, which in some cases result in more severe clinical outcomes in COVID-19 patients^30^. Thus, another requirement for effective surveillance machinery is the ability to distinguish and characterize co-infections, which may complicate consensus genome generation and PANGO lineage determination. After an initial finding of within-host SARS-CoV-2 diversity^31^, reports emerged describing patients likely co-infected with co-circulating SARS-CoV-2 lineages^32–37^. Since our SARS-CoV-2 whole genome sequencing dataset robustly captures these pandemic-wide trends in circulating lineages (**Figure 2**) and is highly stable (**Figure 4**, **Figure 5**), we posited that recovery of multiple SARS-CoV-2 lineage detections from the same sample would be possible.

To identify these potential mixtures (i.e. co-infections), we developed a custom workflow utilizing freyja^14^, an off-the-shelf mixture deconvolution algorithm (Methods). We found that deeply sequenced samples (20.7% or 123,373 of 594,832) produced stable lineage mixture results (>99% genome coverage, 100% S gene coverage, and average depth of coverage > 200X) (**Figure S12a**). We then imposed three criteria for a sample to be classified as a mixture. The first two require the most and second most abundant lineages to have relative abundances no greater than 0.8 and no less than 0.2, respectively, which only 571 (0.46% of 123,373) samples satisfied (**Figure S12b-c**). Thirdly, we required the mixed lineages to differ by at least three defining SNPs (heretofore defined as ‘discriminating SNPs’), since such mixtures were found to have stronger concordance between lineage relative abundances and discriminating SNP allele fractions **Figure S12d-f**). This process yielded a final confident set of 379 mixtures (**Supplementary Data**) likely of similar quality to their non-mixture counterparts, as they were found to have similar median depth of coverage (mixtures: 338.4, non-mixtures: 335.6, p=0.46, Wilcoxon rank-sum test) (**Figure S12g**). These mixtures had similar Ct values as non-mixtures, with the same differences observed between the BA.1 wave samples and those before and afterwards (**Figure S3c**). We also found that these mixtures had lineage compositions concordant with the original lineage called by pangolin^29^, since in most cases the pangolin-derived lineage either closely matched the majority mixture lineage (217 or 57.3%) or was a parent of both majority and minority mixture lineages (142 or 37.5%) (**Figure S13**).

Not surprisingly, we detected a plurality of mixtures during the BA.1 wave (159 or 42%), which represents the largest portion of the dataset analyzed (18% or 22,169 samples). We also observed the highest prevalence of mixtures during the BA.1 wave, peaking at 1.4% the last week of December 2021 (**Figure 6a**). When categorizing the lineages comprised by these mixtures using their lineage groups, we found that most mixtures were of lineages from the same group (347 or 91.6%), e.g. BA.1* mixed with BA.1* (**Figure 6b**, **Table S2**). This is likely due to variants emerging in blocks (**Figure 2**), though we did observe some mixtures of different lineage groups collected during the transitions between these blocks, e.g. one Delta*-Mu* and three BA.1*-Delta* mixtures (**Figure 6b**, **Table S2**).

**Figure 6.**
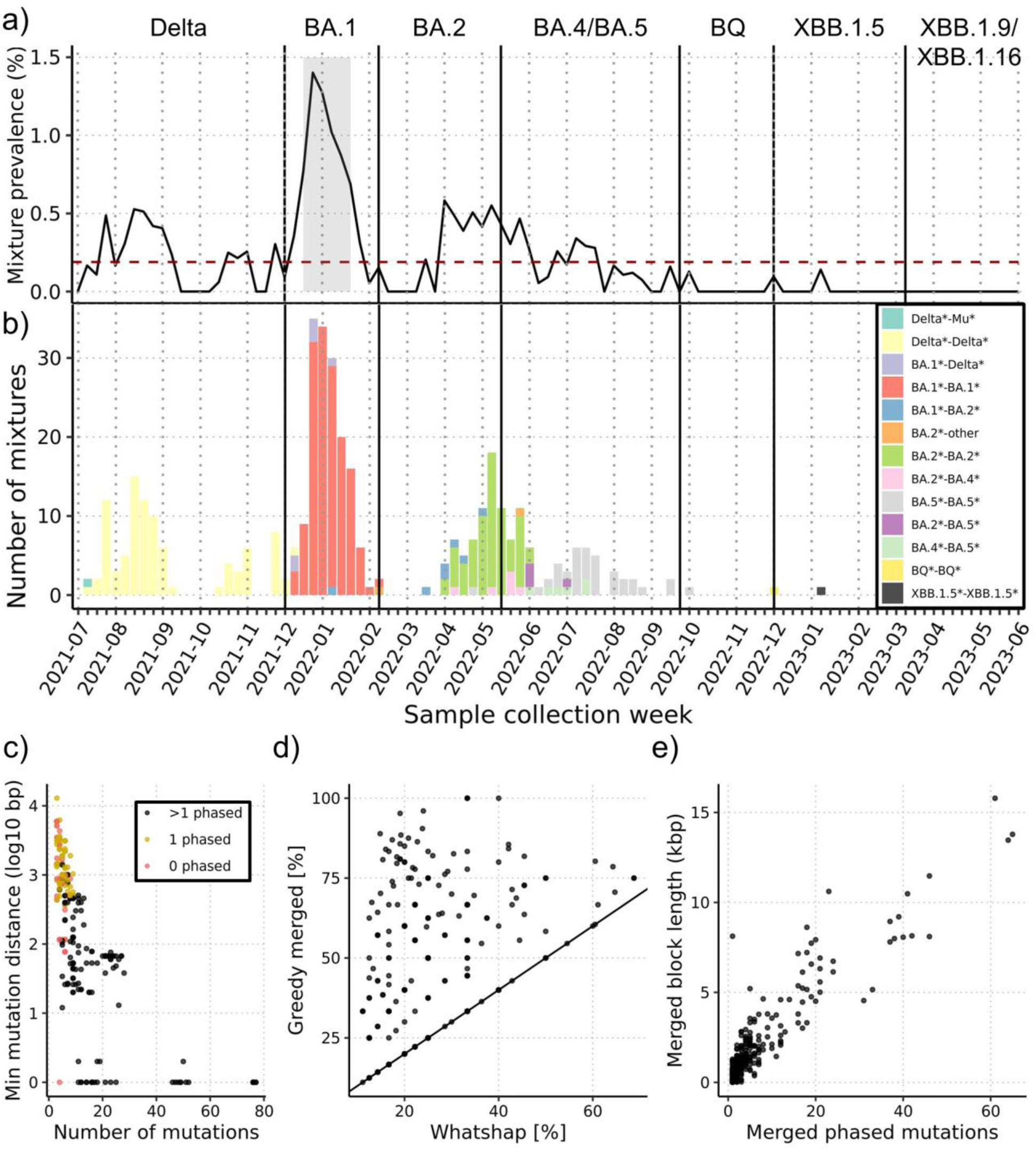
Haplotype phasing of SARS-CoV-2 mixture samples (i.e. coinfections). **a-b)** Prevalence and number of mixtures in each sample collection week with major pandemic waves demarcated as they were in **Figure 3**. In **(a)**, the weekly average (∼0.2%) is shown as a horizontal dashed red line. In **(b)**, mixtures are colored based on the lineage family of the major and minor mixture components, e.g. BA.1*-Delta indicates a mixture of BA.1 and Delta sublineages with majority BA.1. **c)** Sample-level comparison of the number of and minimum distance between UShER defining mutations that discriminate the two mixture lineages. Samples are colored based on their number of defining mutations phased: >1 phased (black, n=149), 1 phased only with non-defining mutation(s) (yellow, n=128), and 0 phased (red, n=102). **d)** Comparison of sample defining mutation phasing among largest resolved haplotype blocks. Results are compared between original whatshap haplotype blocks and those same haplotype blocks merged using freyja mixture results. **e)** The number of defining mutations phased and the lengths of sample merged haplotype blocks.

Since these mixtures have robust sequencing depth (**Figure S12g**) and comprise lineages with as many as 77 discriminating SNPs (**Supplementary Data**), we hypothesized that it would be feasible to resolve lineage haplotypes. Using a standard haplotype phasing tool for long reads (Methods), we were able to produce haplotype blocks in most mixtures (277 or 73.1%), favoring mixtures harboring discriminating SNPs greater in number and closer together (**Figure 6c**). We also developed a custom approach that employs a greedy strategy to merge haplotype blocks together based on the alleles of the discriminating SNPs in each haplotype block (Methods). This greatly increased haplotype block resolution, on average increasing the number of SNPs in the largest (merged) haplotype block by ∼100% and increasing the size of haplotype blocks to as long as 15.8kbp (**Figure 6d-e**). These exceptionally large haplotype blocks were found among the BA.1*/Delta* mixtures, which have the largest number of discriminating SNPs among the mixtures identified (**Supplementary Data**). One example is LC0471172, which is a mixture of BA.1.1.18 and AY.39 that had a final merged haplotype block of length 15.8kbp harboring 61 SNPs and spanning most of ORFs 1a and 1b as well as the entire S gene and 3’ end of the genome (**Figure S14**). These rare, finely resolved mixture haplotypes are evidence that combining haplotype reconstruction with mixture analysis has potential to unveil unique sample characteristics. This mixture analysis workflow thus provides our surveillance apparatus with the essential ability to detect co-infections and ensures that consensus genomes are correctly reported in such cases.

## Discussion

Genomic surveillance, globally and through our contribution to CDC SPHERES, proved to be critical for monitoring the emergence of highly mutated SARS-CoV-2 variants and their potential influence on disease severity^38^ and the hundreds of vaccine development efforts worldwide (183 in clinical development as of March 30, 2023^39^). In this report we showcased our robust, comprehensive U.S.-based SARS-CoV-2 surveillance network enabled through our infrastructure and sequencing capabilities. Our probe-based tiling of the genome precluded surveillance interruptions, while other amplicon-based assays have required multiple updates^10^. However, while our tiled approach has ensured robust lineage detection so far, there is always a possibility that an emerging novel lineage may introduce mutations that could potentially affect our ability to detect it. To address this uncertainty we routinely assess emerging SARS-CoV-2 lineages using our Virseq assay simulator before they are widely circulating among the U.S. population. These resources ensure that we are prepared to swiftly respond to any sudden and/or large mutations in the SARS-CoV-2 genome.

One key feature of our surveillance network is the rapid sequestration and consolidation of high viral titer samples before sequencing. This strategy could in theory be applied to any infectious agent for which we or others have robust diagnostic assays. In the case of SARS-CoV-2 in this study we apply an N1 Ct value upper limit for sample inclusion to guarantee sufficient genetic material for whole genome sequencing. While necessary for sequencing feasibility, this thresholding may introduce bias in the sample selection and comprehensive lineage coverage. Another important aspect of our surveillance is the dense network of various Labcorp^®^ testing centers throughout the U.S. In this study we that show our SARS-CoV-2 diagnostic samples come from all HHS regions and are sent for whole genome sequencing through our Virseq assay with limited bias. Importantly, this comprehensive demographic coverage is contingent on the availability of samples from our testing centers, which could potentially change due to myriad factors such as local mandates and/or health care coverage.

Our targeted long-read sequencing approach is also equipped for recovery of haplotype-resolved viral genomes. To our knowledge, our study is the first to provide a pandemic-wide, high-resolution evaluation of co-infections at this scale, though there have been other systematic efforts that are smaller^40^ or target specific types of co-infections, e.g. Omicron/Delta^37^. A crucial step in confirming co-infections is the haplotype phasing of observed heterozygous mutations, which is generally limited to long-read sequencing approaches described here with PacBio^®^ sequencing and by others using Oxford Nanopore Technologies^®41^. In our study we observe most co-infections from the same lineage group (e.g. BA.1*/BA.1*), likely representing individuals who were exposed to unique variants in rapid succession rather than those who are chronically ill from a previous infection. These co-infections may not only have clinical relevance^30^, but also represent potential recombination events. For example, the BA.1 wave showed the highest prevalence of co-infections in our dataset and incidentally introduced numerous recombinants, including those formed from Delta and BA.1^42–44^. Ultimately, the use of long reads is uniquely suited for distinguishing co-infections from these recombinant cases as well as other sources of intra-host variation of the virus that have been described^31^.

The surveillance apparatus we describe is not only robust to the pandemic undulations but is also flexible and modular. For example, we recently adapted this workflow to employ an Oxford Nanopore Technologies^®^-based ClearLabs^®^ sequencing approach in lieu of our probe-based long-read (PacBio^®^-based) sequencing approach. This alternative EUA approved pipeline enables rapid turnaround time (∼1 day) and retains the same suite of analytical tools as our primary surveillance apparatus. We have also leveraged our infrastructure to provide additional features beyond monitoring SARS-CoV-2 genetic evolution. Early in the COVID-19 pandemic it became clear that convalescent sera from COVID-19 survivors would be essential for development of antibody therapies^45^. We and others analyzed sera specimens collected from over 3,000 unvaccinated individuals and found that most did not exceed antibody concentrations associated with 90% vaccine efficacy, indicating that vaccination is necessary for maximum protection against SARS-CoV-2 infection^46^. We also use our infrastructure to systematically obtain sera from individuals recently infected with SARS-CoV-2 VOCs as part of multiple efforts to investigate antibody cross-reactivity, which is essential for development of COVID-19 vaccine boosters.

As the threat of COVID-19 wanes and global SARS-CoV-2 surveillance networks scale back, there is a strong need for continued development of rapid response tools. Diagnostics that target multiple pathogens, such as our seasonal respiratory panel^47^, are increasingly useful to this end. These diagnostics may serve as outbreak detection tools when their negativity rate spikes, indicating the emergence of a novel pathogen. Agnostic surveillance techniques, such as those monitoring wastewater via shotgun sequencing^13–15,17^, have already shown promise by detecting Poliovirus Type 2 in New York wastewater^48^. If an outbreak does not immediately attenuate, then our surveillance apparatus described in this study could serve as a model for sustained monitoring of whole genome variations that could impact disease severity, outbreak dynamics, and the efficiency of targeted diagnostic assays.

In this retrospective study, we showcase our unique positioning for rapid development and maintenance of robust pathogen surveillance. Our nationwide surveillance network and its suite of analytical and sequencing components collectively serve as a model for future large-scale surveillance efforts. Looking to the future, it is our mission to stay vigilant and continue refining this model to combat the many emergent infectious diseases posing imminent threats to public health.

## Methods

### Ethical Statement

The use of residual de-identified samples for this study was determined as not a human subject research requiring IRB review.

### SARS-CoV-2 surveillance and whole genome sequencing

Extracted total nucleic acid from positive specimens identified through the Labcorp^®^ FDA EUA approved COVID-19 RT-PCR Test or SARS-CoV-2 & Influenza A/B Assay Test were sequestered and consolidated using a Hamilton Microlab^®^ STAR™ instrument and plate selector app, retaining only positive samples with N1 Ct values less than 31. Sample RNA was reverse transcribed to cDNA and a specially designed SARS-CoV-2 probe set containing ∼1000 tiled Molecular Loop^®^ Loopcap™ Molecular Inversion Probes (MIPS) was used to amplify the cDNA from 99.6% of the SARS-CoV-2 genome with most bases covered by 22 MIPs^19^. The product synthesized in-between the MIPs was enriched and had sample specific molecular barcodes added via amplification for long-read sequencing on a Pacific Biosciences^®^ Sequel II™^49^.

### Sequence quality control and post-processing

After sequencing, circular consensus sequence (CCS) bam files were generated using the PacBio^®^ SMRT LINK™ software v9.0 ccs program^50^ and subsequently demultiplexed using lima with the following parameters: “--min-score-lead -1”, “--min-score 80”, “--window-size-multi 1.1”, “-- neighbors”. Molecular Loop^®^ barcodes were then trimmed by aligning sequences to barcodes using pbmm2 with parameters “--sort” and “--preset HIFI” and custom processing scripts. Final sample fastq files were generated by converting the resulting bam files using BamTools^51^.

Sequence fastq files were analyzed using a genome analysis pipeline implemented in the CLC Genomics Server version 9.1.1^52^. This workflow starts with a sample-level fastq file and uses Minimap2^53^ to align reads to the SARS-CoV-2 reference genome (NCBI GenBank reference NC_045512.2) to generate a bam file of the alignment as well as a VCF file containing the variants called using a custom variant caller in CLC. A consensus sequence for each sample was then generated using VCFCons v8.5.0^54^. When VCFCons calls a nucleotide sequence for genome construction it was required to have least 4 CCS reads covering that base pair and an alternate allele frequency compared to the reference of at least 80%. If the alternate allele frequency was between 20% and 80%, then the appropriate ambiguous IUPAC nucleotide was called. If a nucleotide was covered by less than 4 CCS reads it was reported as ambiguous (N) in the consensus sequence.

Three different coverage quality control metrics were used to ensure high accuracy of resulting consensus genome sequences. Firstly, the median CCS read coverage was calculated separately for 29 ∼1kb genomic regions and each sample was required to have at minimum 10X mean of median amplicon coverage (depth of coverage). For samples to be kept for downstream Phylogenetic Assignment of Named Global Outbreak (PANGO) lineage determination^18^ and sequence analysis, a minimum genome coverage of 50% was required. For sample genome sequences to be reported to the CDC (and later deposited to GISAID^20^), a more stringent genome coverage threshold of 90% was applied along with a third coverage filter that required no more than 1 ambiguous base call in each 6bp sliding window of the S gene. Furthermore, samples were reported within 21 days of collection.

Sample consensus genome sequences with at least 10X depth of coverage and genome coverage of at least 50% were further analyzed using pangolin software^29^ (v4.3.1 with pangolin data v1.22) and the UShER algorithm^55^ with default parameters to determine SARS-CoV-2 PANGO lineages. Sequences and their mutations were also characterized using Nextclade v2.14.0^56^ and the Nextclade SARS-CoV-2 dataset compiled on August 9^th^, 2023. In all subsequent analyses, PANGO lineages were assigned groups by a pre-determined set of parent lineages, representing key variants of concern (VOCs) throughout the pandemic. A lineage and its descendants are indicated by appending “*”. When a lineage is a descendant of two of the following parent lineage groups, the closest parent was selected. If a lineage was not a descendant of any of the parents, then it was placed in the “other” group. Parent lineage groups are as follows: Alpha (B.1.1.7*), Beta (B.1.351*), Gamma (P.1*), Epsilon (B.1.427* and B.1.429*), Eta (B.1.525*), Iota (B.1.526*), Kappa (B.1.617.1*), Mu (B.1.621*), Zeta (P.2*), Delta (B.1.617.2*), BA.1*, BA.2*, BA.4*, BA.5*, BQ*, XBB*, XBB.1.5*, XBB.1.9*, and XBB.1.16*.

### Demographic and phylogenetic analysis

Time-resolved phylogenetic analysis of SARS-CoV-2 consensus genome sequences was performed using augur v.22.2.0^57^ and auspice.us v0.12.0 (Auspice 2.49.0) within the Nextstrain framework^58^. Consensus genomes were restricted to those with 100% S gene coverage and at least 99% genome coverage sequenced between January 2021 and June 2023. The augur filter utility was used to limit the dataset to a maximum of 1,000 sequences per month each with metadata including sample collection date and PANGO lineage. Next, the augur refine utility was used to create a time-resolved phylogenetic tree using the TreeTime algorithm^59^. Finally, the resulting tree was annotated with lineage information using the augur export utility, and the final tree was displayed on auspice.us.

Age and gender distributions of all samples in this study with consensus genomes passing the minimal coverage criteria for PANGO lineage determination were analyzed, restricting to those collected between January 2021 and June 2023 with weekly sample counts of at least 100. Distributions were also stratified by U.S. HHS regions^60^. For each region, annual gender proportions were calculated, and age distributions were determined by aggregating ages into different groups as follows: 0-17 (pediatrics), 18-19, 20-29, 30-39, 40-49, 50-59, 60-69, 70-79, and 80 or older.

### RT-PCR sampling, positivity, and N1 Ct value analysis

Demographic data and positive/negative results of RT-PCR samples collected through the surveillance network described in this study were aggregated for multiple analyses. First, the RT-PCR sampling was compared with Virseq assay sampling across each HHS region and month where at least 100 Virseq samples were sequenced. The geographic distribution of RT-PCR and Virseq sampling were also independently assessed by normalizing with U.S. state census data from 2020-2022^61^. Census data from 2023 was estimated by averaging that from 2020-2022. Overall RT-PCR positivity was analyzed weekly and was also stratified by gender and age groups (described in Methods section ‘Demographic and phylogenetic analysis’). N1 Ct values of Virseq samples were analyzed weekly and stratified based on whether samples were collected before or after the Omicron wave (November 2021 to February 2022), denoted as ‘pre-wave’, ‘Omicron wave’, and ‘post-wave’. N1 Ct values were also compared based on co-infection status using only deeply sequenced samples from the co-infection analysis described later in Methods section ‘Workflow for detection and characterization of SARS-CoV-2 mixtures’.

### Whole genome coverage and mutation frequency analysis

Genome-wide assessment of SARS-CoV-2 sequence mutation frequencies was performed using all results obtained from Nextclade v2.14.0^56^ for samples passing minimal coverage quality control. Mutations were considered as “lineage-defining” if they appeared in at least 70% of the genome sequences assigned to that lineage. The number of mutations with at least 5% prevalence across the entire genome and within specific SARS-CoV-2 genic regions was calculated for each sample collection week. Circulating lineage diversity was calculated in each week using unique lineage counts and the Shannon entropy implementation in the vegan R package^62^. In all analyses that indicate dominant lineage groups during specific time periods, this was calculated using the first week where at least 5% of the lineages were assigned to that lineage group. In the case of XBB.1.9 and XBB.1.16, these were combined, and the sum of their occurrences was used.

Per-base CCS coverage analysis was performed using selected VOCs (B.1.1.7, B.1.617.2, BA.1, BA.2, BA.4, BA.5, BQ.1, and XBB.1.5), where the sequences chosen were required to have depth of coverages of 100 +/- 10. 50 sequences were randomly chosen for each VOC that met the coverage constraint and had the exact PANGO lineage determination of the VOC. Per-base coverage was determined using Samtools^63^ depth with parameters “-q 0 -Q 0” applied to the sample alignment bam files. Median per-base coverage was then calculated for each VOC and smoothed using a 30bp sliding window. Lineage defining mutation density was also determined for each VOC by enumerating mutations at each genomic position and smoothing over a 1kbp sliding window.

### Virseq performance simulator

The Virseq pipeline performance was assessed by constructing a process whereby an input whole genome sequence is mutated in a manner that simulates the coverage and errors introduced by sequencing and post-processing analysis. This simulator was constructed using coverage and error models of a representative sequencing batch selected from February 2022 containing 520 samples reported to CDC (**Supplementary Data**). The coverage model was designed using a min max normalization strategy, where two components were stored for later application of the model: 1) a list of each sample’s maximum per-base coverage and 2) the mean min max normalized coverage at each position of the genome. In practice, since the peripheral 5’ and 3’ ends of the genome do not have tiled coverage by design, a sample’s min max normalized coverages equate to positional coverage divided by the maximum per-base coverage. The error model was generated under the assumption that the consensus at each base position is the correct base call. Thus, the probability of an error was calculated using the maximum minor allele frequency. At each genomic position the number of each base call was enumerated in R using Rsamtools^64^ to identify the error rates for each sample, and the mean error rate at each position was recorded. If the median coverage at a position was less than 3, then the global median error rate was used. If the error rate was zero (i.e. there were not any alternate base calls at a position), then half of the global minimum non-zero error rate was used instead.

These two models were collectively applied to an input SARS-CoV-2 whole genome sequence first by identifying positional coverages. A maximum coverage was randomly selected from the list obtained from the representative batch and the expected mean coverage was computed at each position by multiplying this maximum coverage by each pre-computed mean positional fractional coverage. Next, the coverage at a position was sampled from a Poisson distribution using the mean obtained in the prior step. If the sampled coverage was less than the minimum per-base coverage threshold of 4, then an ambiguous base call was simulated at that position (see Methods section ‘Sequence quality control and post-processing’). Next, for all remaining unambiguous positions a consensus base call was simulated using the sampled coverage and the pre-computed average error at each position. Bases were sampled using a cumulative binomial using the average error and coverage, and if the number of errors exceeded half of the base calls a random base was called at the position; otherwise, the reference base was used. Finally, the PANGO lineages were determined for both the original and simulated sequence using pangolin software^29^ (v4.3.1 with pangolin data v1.22) and checked for concordance. If the lineages were identical, the result was considered an ‘exact match’. If one lineage was a descendant of the other, then the result was categorized as a ‘parent match’. All other cases were considered ‘discordant’.

Two simulation experiments were used to assess the performance of the Virseq pipeline. Sequences were retrieved from GISAID^20^ (accessed August 25^th^, 2023) and those used in the experiments were restricted to those with at least 99% genome coverage, ensuring high quality lineage calls. The first experiment assessed up to 100 sequences from each VOC and descendant lineages thereof, current and former, designated in pangolin data v1.22. The second experiment used a random selection of 10,000 sequences from each month ranging from January 2021 to July 2023.

### Workflow for detection and characterization of SARS-CoV-2 mixtures

A custom workflow was developed to detect and characterize samples containing more than one unique PANGO lineage, i.e. mixtures or co-infections. All bam files of samples passing minimum coverage metrics were processed through the recommended freyja^14^ processing workflow, first calling variants using iVar^65^ and subsequently using the freyja demixing algorithm^14^ to determine the lineage abundances in each sample. This algorithm attempts to identify a parsimonious set of lineages best explaining the UShER^55^ defining single nucleotide polymorphisms (SNPs) detected in the sample.

Lineage abundances were then processed, greedily aggregating abundances of lineages with parent/descendant relationships (e.g. BA.1 and BA.1.1) starting with the most abundant lineage. This was performed since one lineage would have UShER^55^ defining SNPs forming a subset or superset of the other lineage, and it was assumed that this splitting of highly similar lineages was due to sequencing noise. Samples were then filtered, requiring an empirically determined minimum depth of coverage where the rate of mixture detection was stable and low (**Supplementary** Figure 12a). Mixtures were then required to satisfy three criteria: 1) Top lineage relative abundance no greater than 0.8, 2) second lineage relative abundance no less than 0.2, and 3) minimum of 3 UShER^55^ defining SNPs discriminating the two lineages comprising the mixture (**Supplementary** Figure 12b-f). Each resulting mixture sample was then categorized using the parent lineage groups of the two mixed lineages (see Methods section ‘Sequence quality control and post-processing’).

Mixture samples were then processed using WhatsHap^66^, a standard haplotype assembly tool suitable for long sequencing reads. If a sample did not yield any haplotype blocks (i.e. no two SNPs were phased), then analysis was halted. If only a single haplotype block was obtained, then the block length and phased mutations therein were recorded. If a sample had at least two haplotype blocks, then a greedy algorithm was applied to merge these blocks while leveraging *a priori* mixture knowledge yielded by freyja^14^. The number of UShER^55^ defining SNPs unique to each mixture lineage (i.e. lineage-discriminating SNPs) was recorded for each block, and blocks were iteratively merged in order of descending number of lineage-discriminating SNPs (ties broken by using the larger block length). Blocks were merged to maximize the number of correctly phased lineage-discriminating SNPs. If the addition of a block to this greedily merged block didn’t improve this optimization criterion, then it was skipped, and the next block was assessed. This process continued until no further blocks with lineage-discriminating SNPs remained. A custom script was used to modify the haplotagged bam output by WhatsHap^66^ for visualization of merged haplotype blocks in Integrative Genomics Viewer v2.16.2^67^.

## Author Contributions

Development and maintenance of Virseq surveillance apparatus – all authors. Manuscript supervision and administration – L.K.I. Manuscript conceptualization and methodology – H.N.B., K.S., Q.Zh., J.D.W., S.L., and L.K.I. Formal analysis and data curation – H.N.B. and K.S. Formal analysis review – H.N.B., K.S., Q.Zh., S.L., and L.K.I. Preparation of manuscript figures and tables – H.N.B. and K.S. Original manuscript draft preparation – H.N.B. Manuscript review and editing – H.N.B., K.S., Q.Zh., Q.Ze., J.D.W., M.B.N., S.E.D., J.Me., M.E., S.L., and L.K.I.

Approval of final manuscript – all authors.

## Funding Statement

No grants or financial support were used for this study.

## Acknowledgements

We would like to acknowledge the dedicated members of our IT infrastructure team who continue to be instrumental in maintaining the resources needed for this surveillance effort. We would also like to thank the hundreds of technicians and technologists who processed SARS-CoV-2 PCR testing at Labcorp during the pandemic. Without the hard work and dedication of the broader Labcorp enterprise, this surveillance effort would not have been possible. We would also like to extend our gratitude to the dedicated teams at PacBio and Molecular Loop who were instrumental in helping us set up their respective resources for use in the Virseq pipeline. We would especially like to thank Elizabeth Tseng at PacBio for her significant contribution towards the development of the VCFCons software used in the Virseq pipeline. Lastly, we would like to thank the U.S. Centers for Disease Control (CDC) for their guidance throughout this surveillance effort.

## Competing Interests

All authors are current or former employees of Labcorp, a provider of clinical diagnostic services.

## Data Availability

**Figures S1-13** and **Table S1** may be found in the **Supplementary Materials**. A list of de-identified LCIDs and their corresponding GISAID EPI_ISL IDs from samples analyzed in this study that were reported to CDC may be found in the **Supplementary Data**. A list of LCIDs used to construct the empirical simulation model are included in the **Supplementary Data**. Additionally, all processed data used to generate Figures 2b, **3b**, **4-6**, and **S1-13** are provided in the **Supplementary Data**. Data used to generate Figure 3a are provided in **Table S1**.

## Notes

### Author Declarations

Western Institutional Review Board waived ethical approval for this work.

### Summary of Updates

Author list and affiliations updated.

