## Supplementary Materials for "A program for real-time surveillance of SARS-CoV-2 genetics"

Hayden N. Brochu, *et al*


| Page # | Supplementary Figure/Table |
|---|---|
| 2 | Figure S1. SARS-CoV-2 PANGO lineage analysis of high-quality genomes from USA samples collected between 2021 January and 2023 July in HHS regions 1-5. |
| 3 | Figure S2. SARS-CoV-2 PANGO lineage analysis of high-quality genomes from USA samples collected between 2021 January and 2023 July in HHS regions 6-10. |
| 4 | Figure S3. Analysis of diagnostic qPCR N1 Ct values. |
| 5 | Figure S4. Comparison of RT-PCR and Virseq sampling stratified by HHS regions. |
| 6 | Figure S5. Age and gender proportions of USA samples collected between January 2021 and July 2023 in HHS regions 1-5. |
| 7 | Figure S6. Age and gender proportions of USA samples collected between January 2021 and July 2023 in HHS regions 6-10. |
| 8 | Figure S7. Trends in PCR positivity across patient age and sex. |
| 9 | Figure S8. Robustness of whole genome probe-based sequencing with zero mutation tolerance. |
| 10 | Figure S9. Genome-wide mutations and per-base coverage of common lineages detected throughout the SARS-CoV-2 pandemic. |
| 11 | Figure S10. Virseq performance simulator error and coverage models. |
| 12 | Figure S11. Coverage and error results from Virseq simulations. |
| 13-14 | Figure S12. Curation of high-quality SARS-CoV-2 mixture samples. |
| 15 | Figure S13. Concordance between PANGO lineage calls determined by pangolin and the majority/minority mixture lineages determined by freyja. |
| 16 | Figure S14. Integrative Genomics Viewer depiction of sample LC0471172, which had the largest merged haplotype block size (~15.8 kbp). |
| 17 | Table S1. Raw and log10-transformed sample counts for each U.S. state and region (District of Columbia). |
| 18 | Table S2. SARS-CoV-2 mixture lineage group co-occurrence frequencies. Each entry in the table represents the frequency of two lineage groups co-occurring in a sample, where the most and second-most abundant lineage groups are shown on the left and top, respectively. |

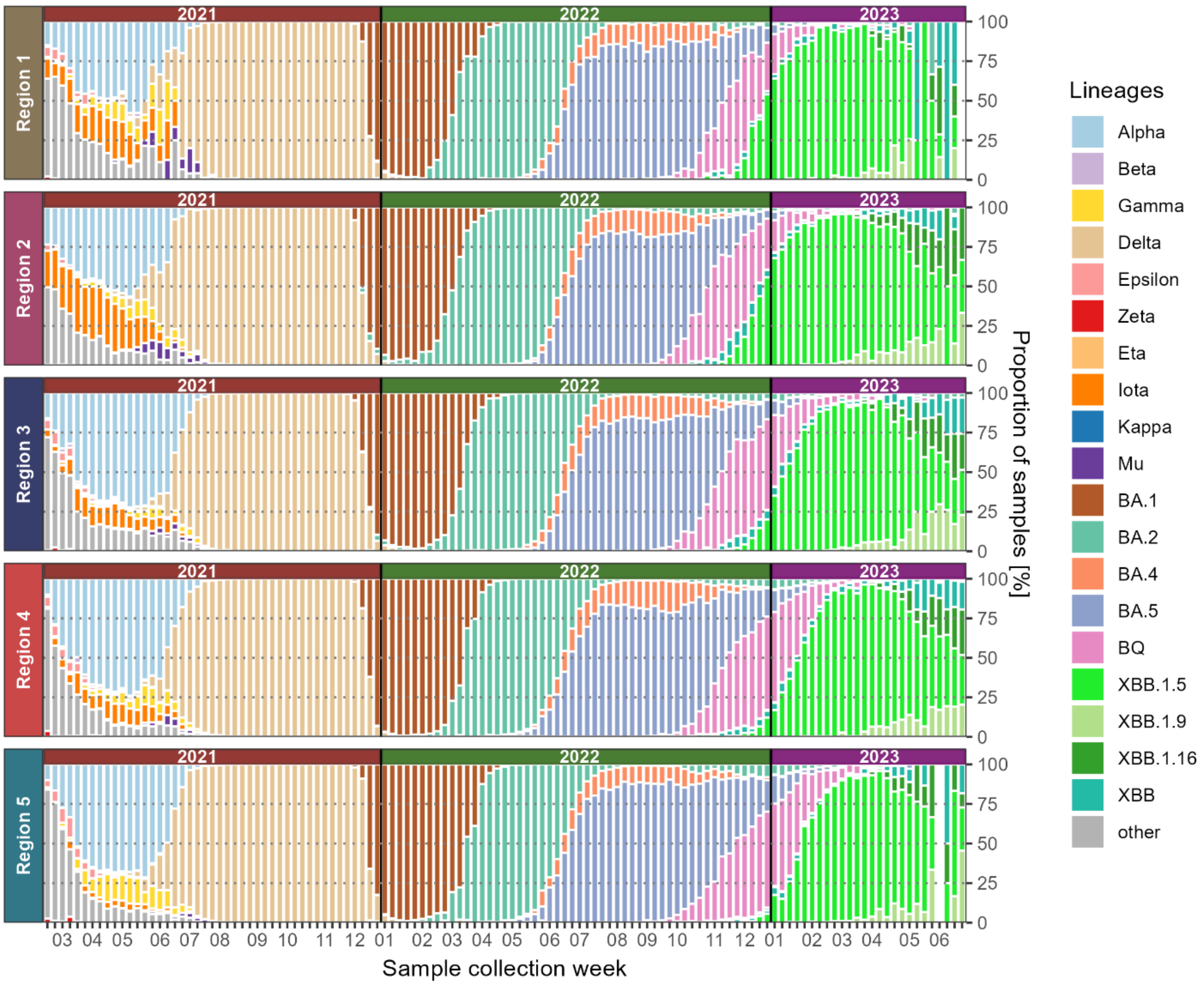


**Figure S1. SARS-CoV-2 PANGO lineage analysis of high-quality genomes from USA samples collected between 2021 January and 2023 July in HHS regions 1-5.** Samples are stratified by U.S. HHS regions, labelled on the left of each plot.

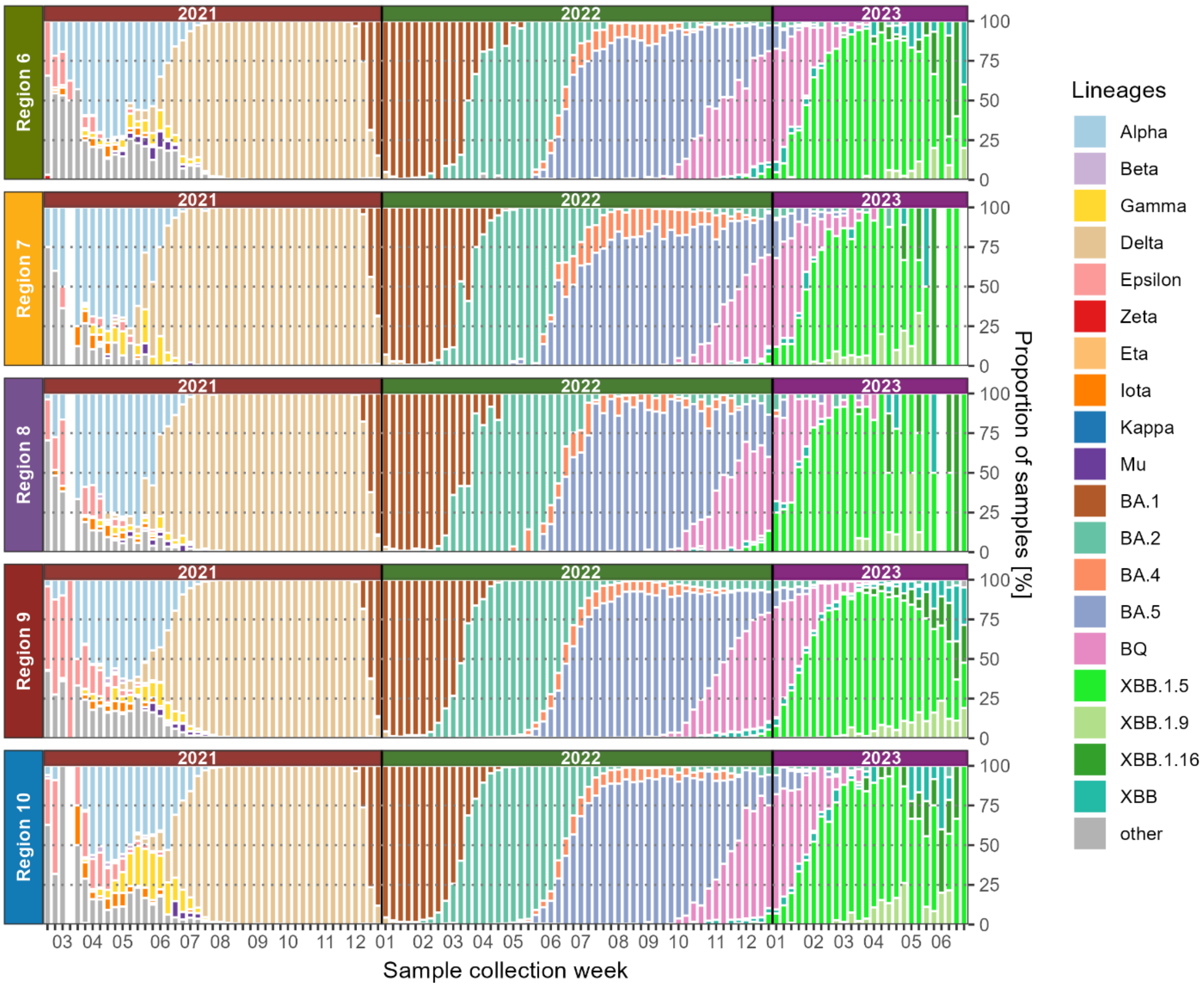


**Figure S2. SARS-CoV-2 PANGO lineage analysis of high-quality genomes from USA samples collected between 2021 January and 2023 July in HHS regions 6-10.** Samples are stratified by U.S. HHS regions, labelled on the left of each plot.

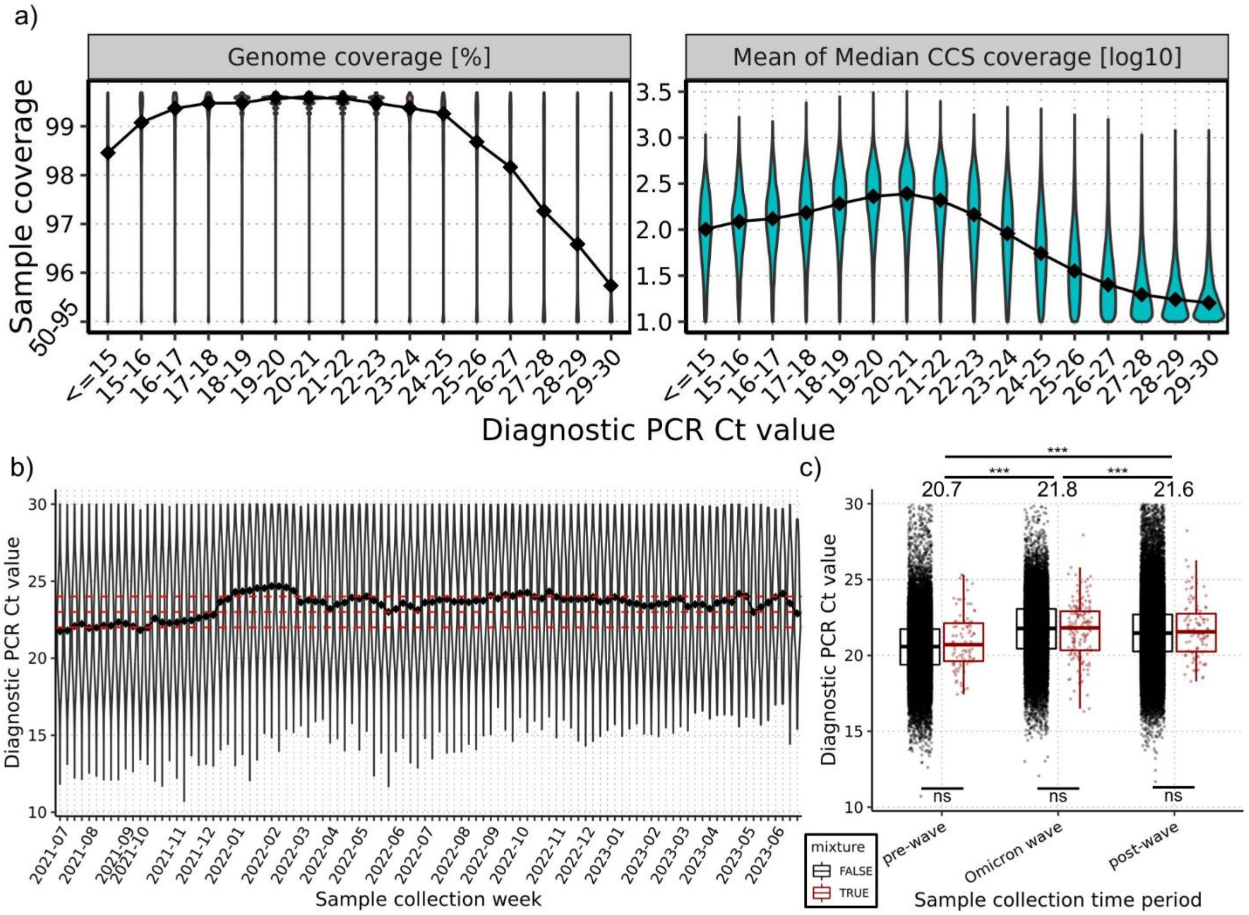


**Figure S3. Analysis of diagnostic qPCR N1 Ct values. a)** Distribution of sample coverages (left: genome coverage [%], right: Mean of median CCS coverage [log10]) stratified by their Ct values. Curves and data points represent the medians of each distribution. **b)** Distribution of Ct values stratified by sample collection week, with curve and data points representing the medians of each distribution. Horizontal red dashed lines indicate Ct values 22, 23, and 24, bottom to top. **c)** Distribution of Ct values stratified by period of the pandemic: Omicron wave (11/2021-2/2022), pre-wave, and post-wave. These distributions are stratified by whether samples were identified as mixtures (red) or not (black) and are restricted to those with high coverage (mean of median CCS coverage >= 200) used in the mixture analysis. Median Ct values and statistical comparisons of the combined distributions from each period (black and red, combined) are shown above the plot. Statistical comparisons based on mixture status are shown below the plot without medians shown, as they were identical in all comparisons, matching the value shown above the plot. Statistical significance was calculated using Wilcoxon rank-sum tests; ***p<0.001, ns=not significant.

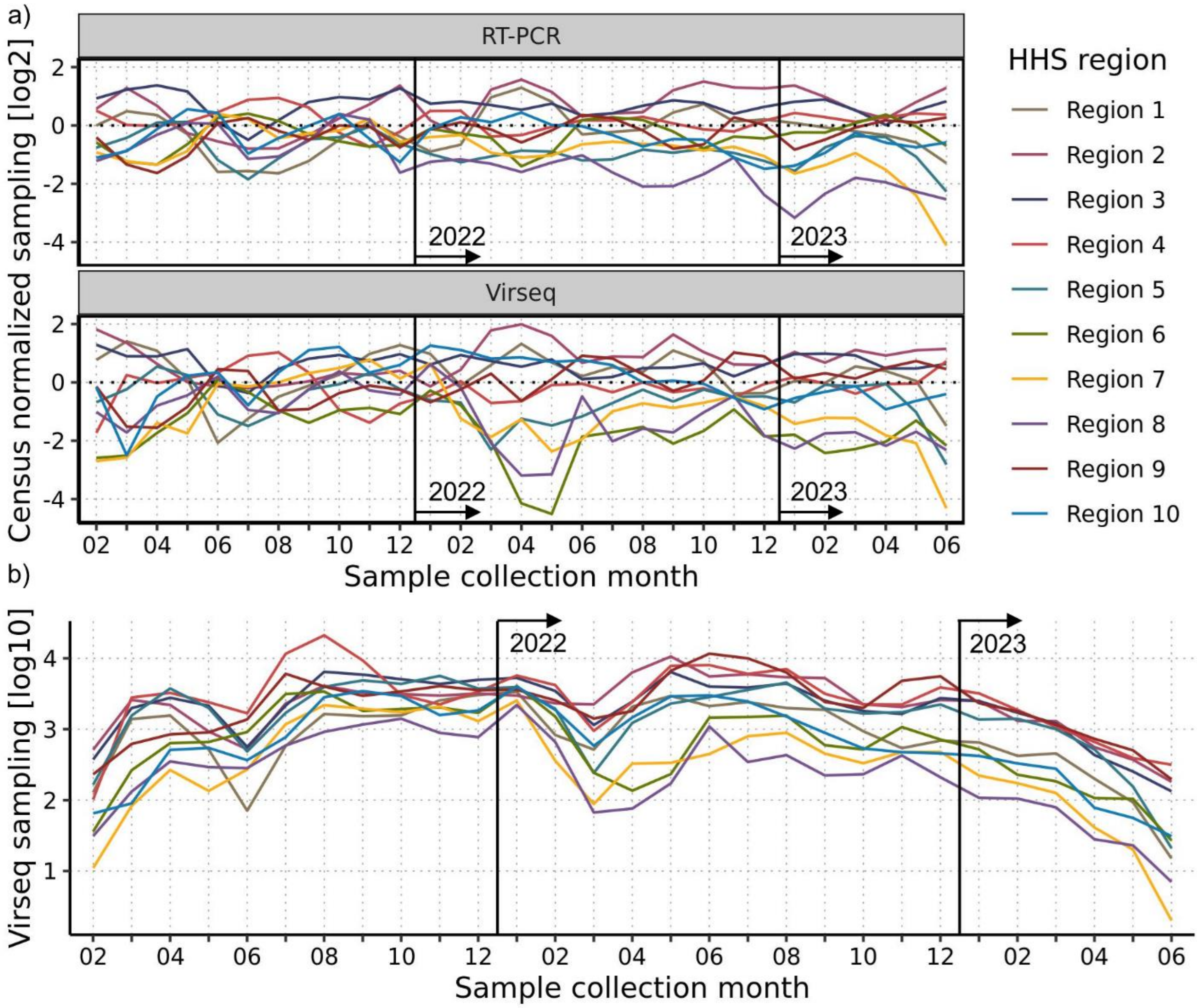


**Figure S4. Comparison of RT-PCR and Virseq sampling stratified by HHS regions. a)** Census normalized monthly sampling (log2) of RT-PCR (top) and Virseq (bottom) sampling. **b)** Total monthly Virseq sampling (log10). In both panels HHS regions are represented by different colored lines and years are separated by vertical black lines with the new year marked to the right of the lines.

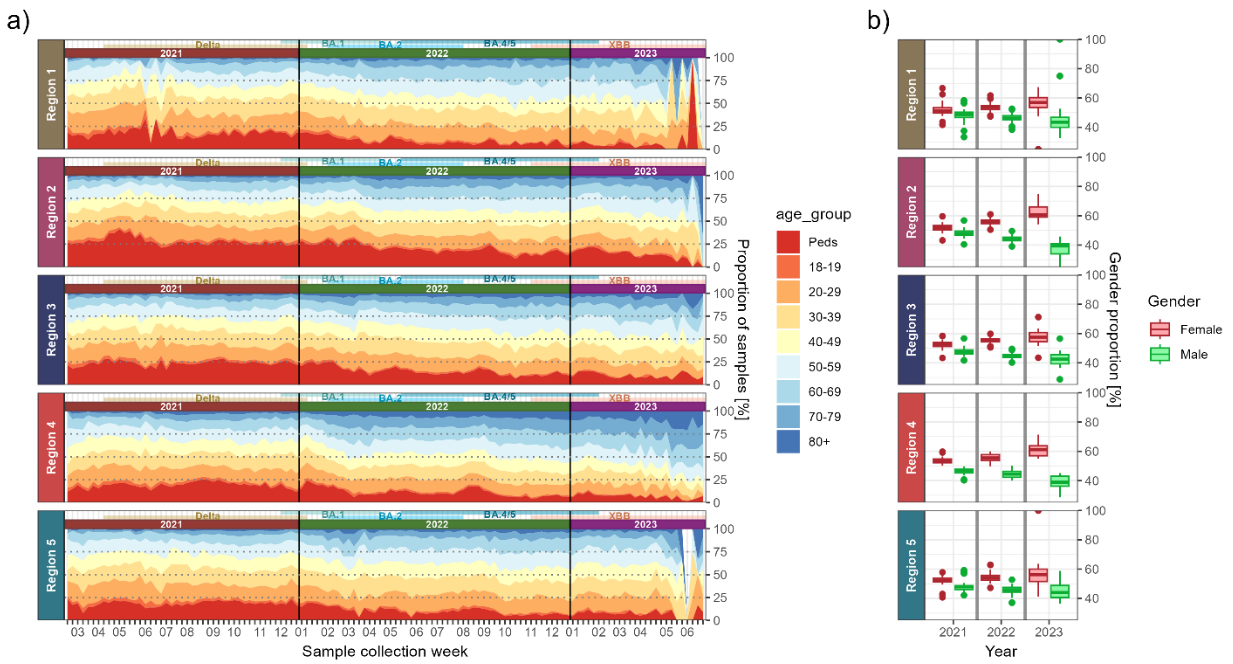


**Figure S5. Age and gender proportions of USA samples collected between January 2021 and July 2023 in HHS regions 1-5.** Each plot is stratified by U.S. HHS region, labelled on the left of each plot. **a)** Weekly age distributions are shown with age group proportions in different colors and the dominant lineages (prevalence > 5%) from different time periods of the pandemic shown above each panel. **b)** Weekly female and male proportions stratified by year (2021, 2022, and 2023) with females and males colored in red and green, respectively.

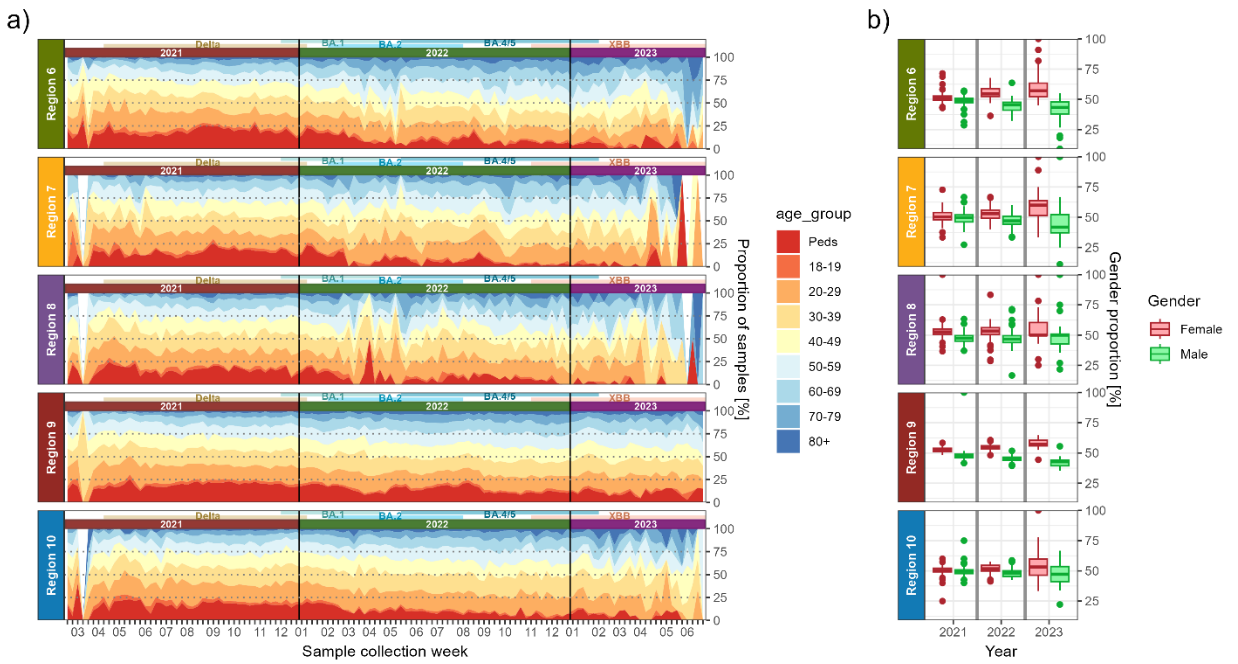


**Figure S6. Age and gender proportions of USA samples collected between January 2021 and July 2023 in HHS regions 6-10.** Each plot is stratified by U.S. HHS region, labelled on the left of each plot. **a)** Weekly age distributions are shown with age group proportions in different colors and the dominant lineages (prevalence > 5%) from different time periods of the pandemic shown above each panel. **b)** Weekly female and male proportions stratified by year (2021, 2022, and 2023) with females and males colored in red and green, respectively.

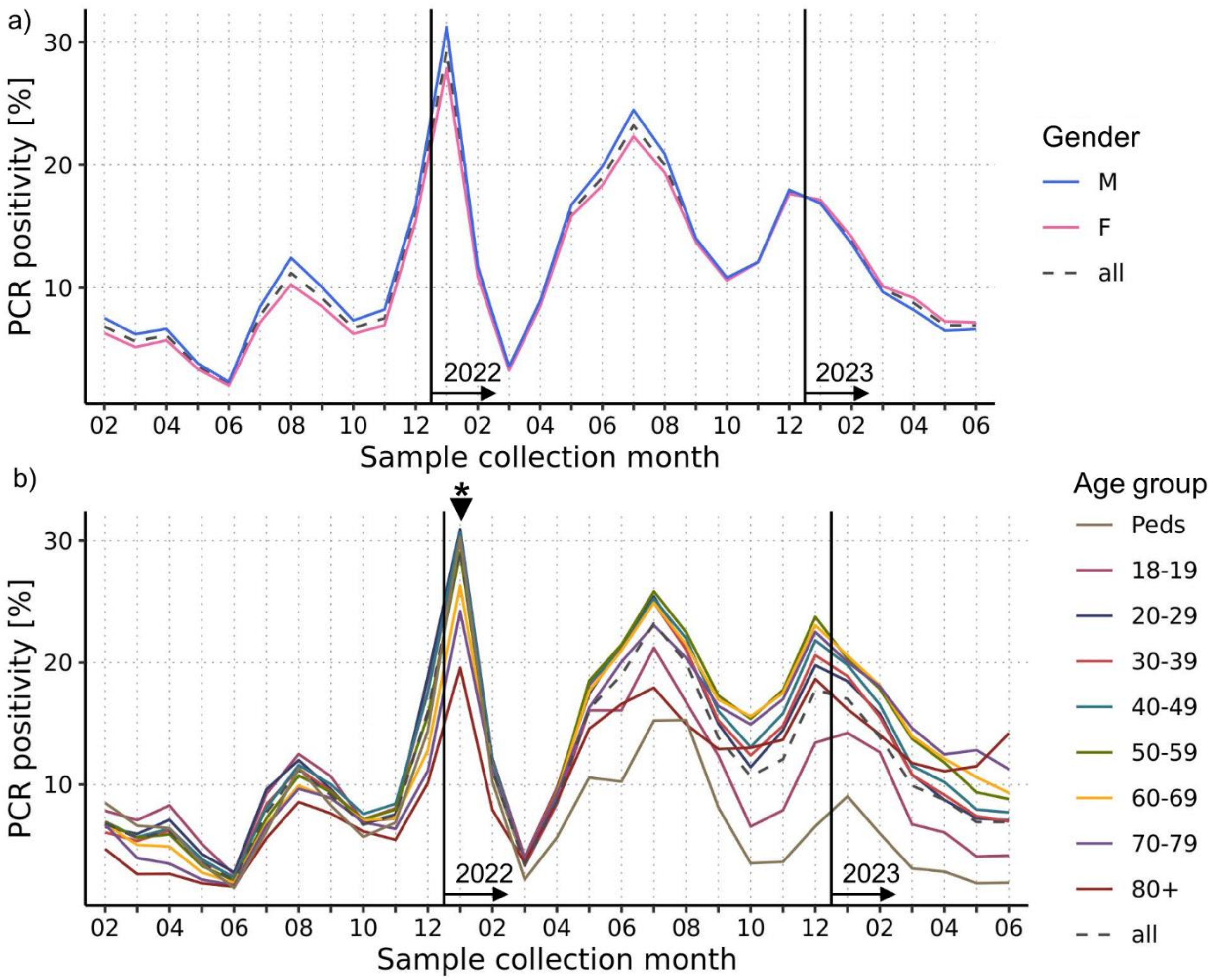


**Figure S7. Trends in PCR positivity across patient age and sex.** Average PCR positivity is shown for each sample collection month, stratifying by patient gender **(a)** and age group **(b)** in different colored lines. The overall positivity is shown as a dashed black line in each panel. Years are separated by vertical black lines and the new year is marked to the right at the bottom of each line. In **(b)**, a small arrow and asterisk is notated at the top above January 2022, signifying multiple announcements by the FDA and CDC pertaining to booster and primary COVID-19 vaccines for adolescents.

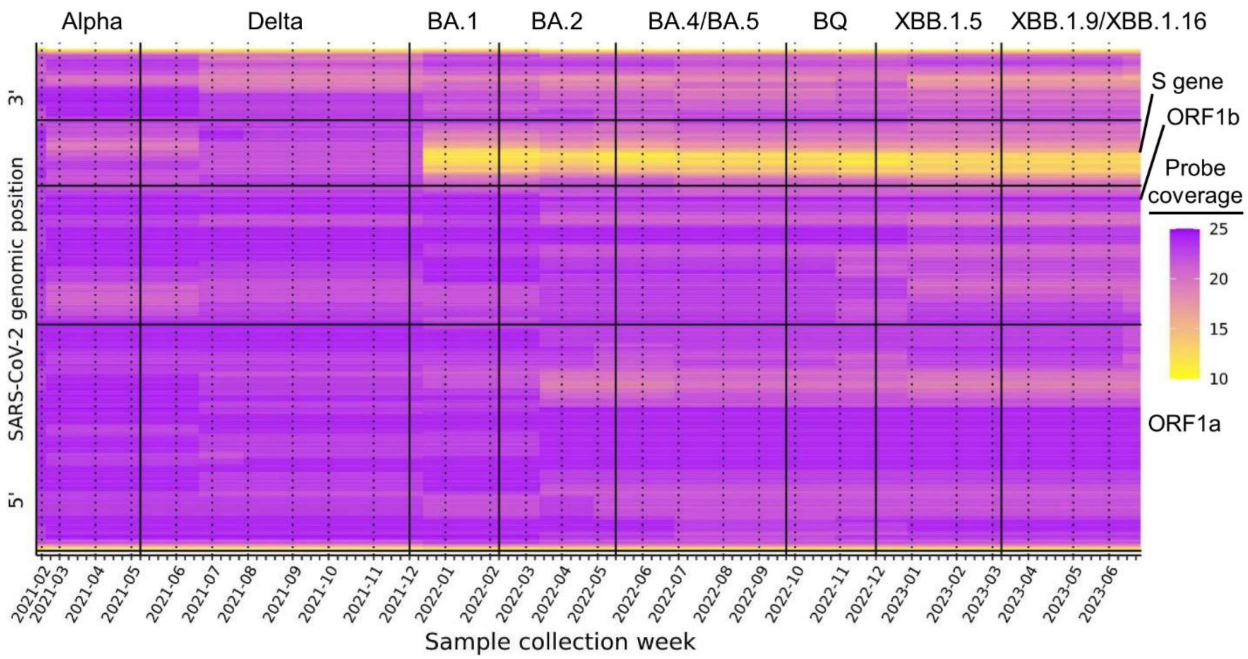


**Figure S8. Robustness of whole genome probe-based sequencing with zero mutation tolerance.** Heatmap showing the genome-wide probe coverage of the most common lineage in circulation for each collection week with genomic positions shown 5' (bottom) to 3' (top). Probes were considered failures if a deletion, insertion, or SNP was detected in either probe arm. Large genic regions (ORF1a, ORF1b, S) are indicated by horizontal lines and are labeled on the right. Results are stratified by sample collection week with vertical bars separating the major waves of the pandemic with the causal variant shown above. Waves are demarcated using the collection week when the causal variant first reached 5% prevalence.

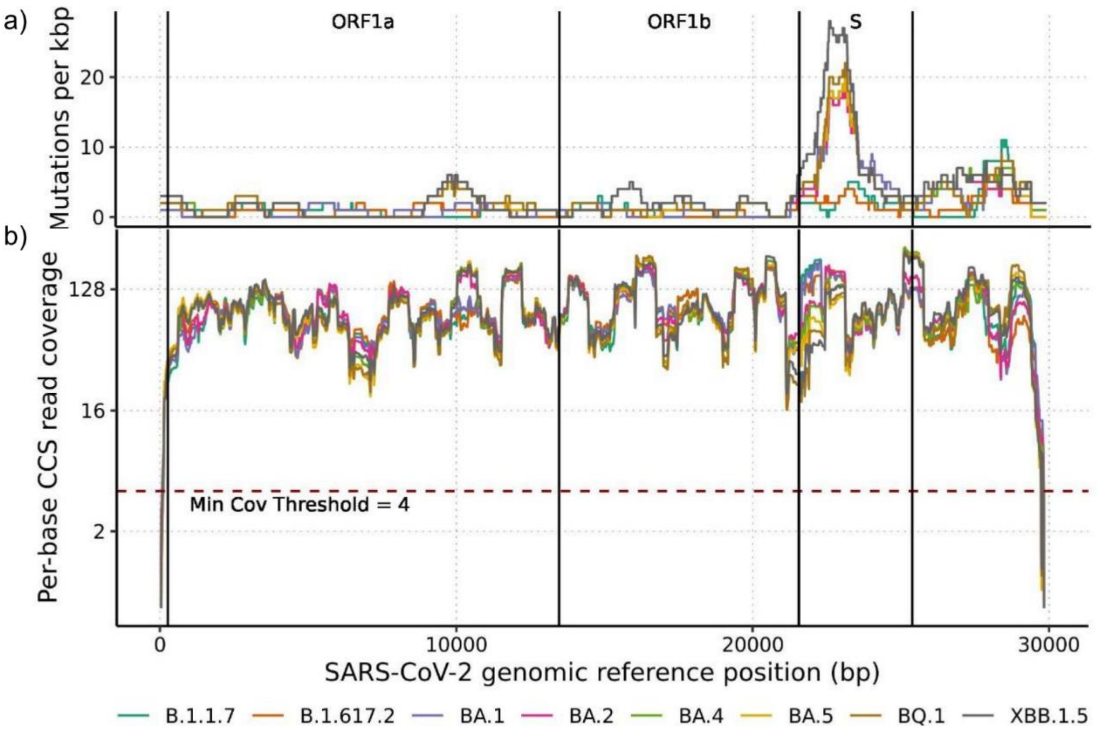


**Figure S9. Genome-wide mutations and per-base coverage of common lineages detected throughout the SARS-CoV-2 pandemic. a)** Number of mutations are shown over 1 kbp sliding windows. **b)** Per-base CCS read coverage (log2 scale) is shown with the minimum coverage threshold of 4 shown with a horizontal dashed red line. In both panels, major genic regions are demarcated by vertical lines with labels shown above and SARS-CoV-2 genomic positions are shown 5' (left) to 3' (right).

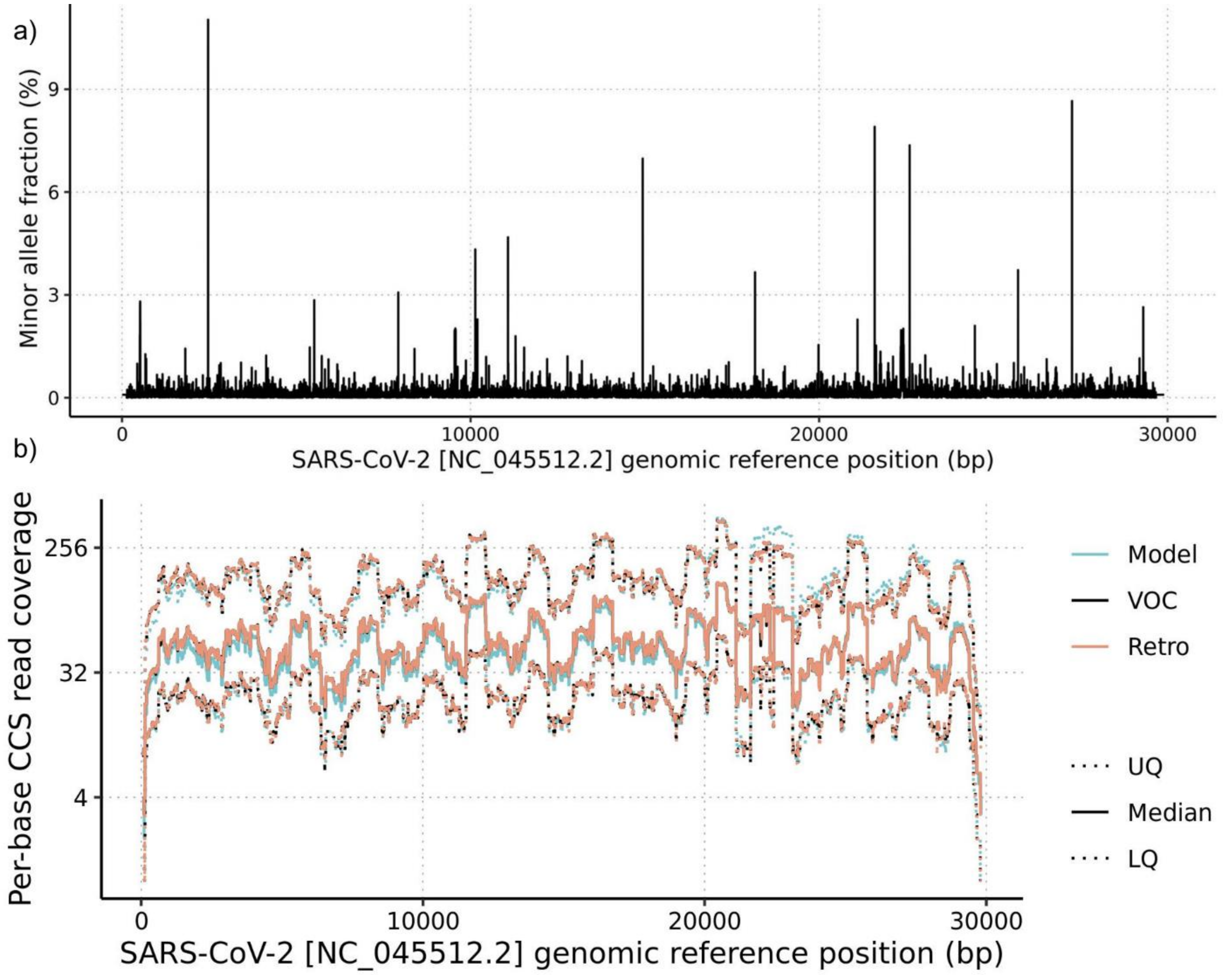


**Figure S10. Virseq performance simulator error and coverage models. a)** Mean minor allele fraction (%) at each position of the SARS-CoV-2 reference genome, representing the error model. **b)** Per-base CCS read coverage distribution of samples used to generate the coverage model (blue) as well as 10,000 randomly chosen samples each from the retrospective (Retro, red) and VOC (black) simulation experiments. The lower quartile (LQ) and upper quartile (UQ) are shown as dotted lines, and the median is shown as a solid line.

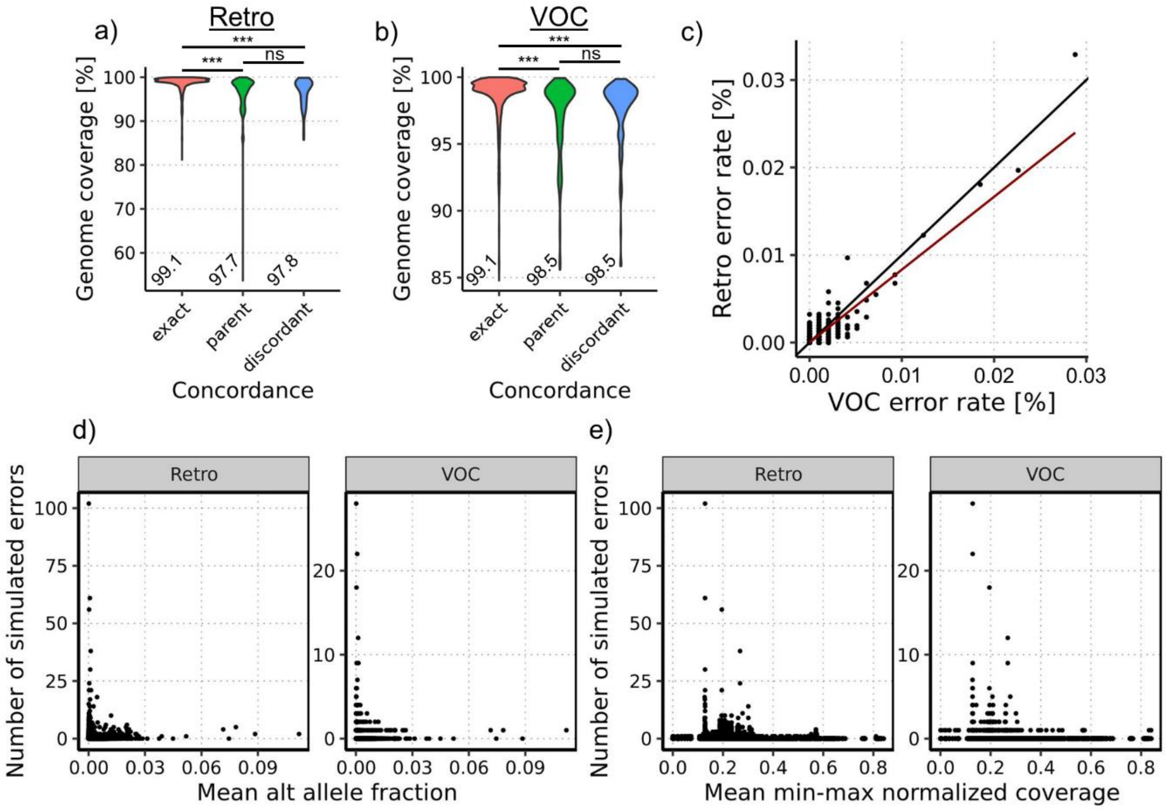


**Figure S11. Coverage and error results from Virseq simulations.** Genome coverage (%) violin plots are shown for the retrospective (retro) **(a)** and VOC **(b)** experiments, stratified by the PANGO lineage concordance result of the simulated genome with median coverages shown below the plotted distributions. **c)** Scatter plot comparing the error rates at each genomic position across all genomes simulated for the retrospective (y-axis) and VOC (x-axis) experiments, with the red and black lines indicating a simple linear fit and y=x line, respectively. The number of simulated errors generated at each genomic position is shown for the retrospective and VOC experiments compared against the **(d)** mean alternate allele fraction and the **(e)** mean min-max normalized coverage. Statistical comparisons of coverage distributions performed using Wilcoxon rank-sum tests; ***p<0.001, ns = not significant.

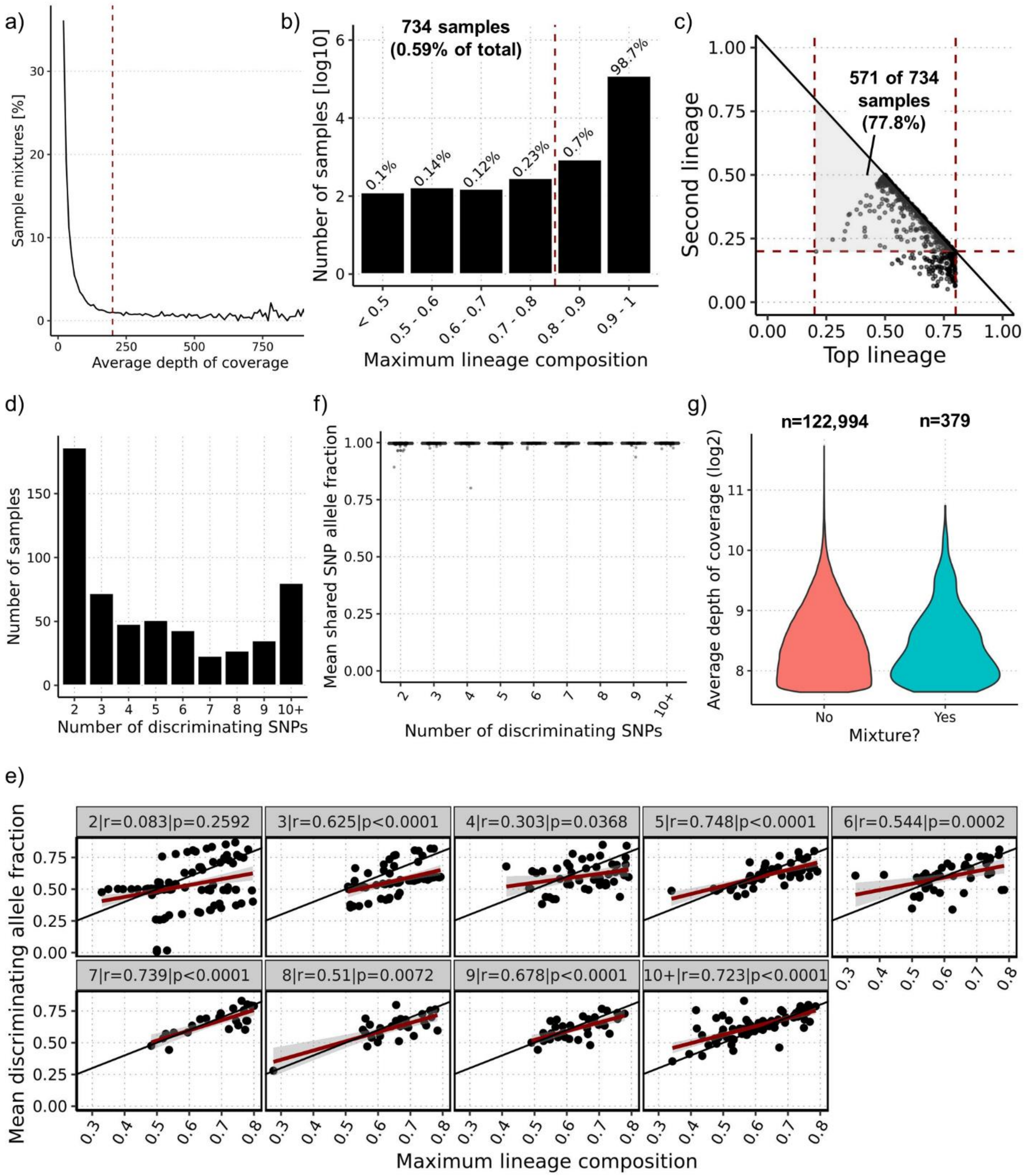


**Figure S12. Curation of high-quality SARS-CoV-2 mixture samples. a)** Percentage of samples initially flagged as mixtures (maximum lineage composition < 0.8) when imposing increasingly stringent coverage thresholds for samples to be included in mixture analysis. The vertical red dashed line indicates the coverage threshold chosen (200). **b)** Distribution of samples binned by maximum lineage composition with the vertical dashed line indicating the threshold under which a sample was initially considered a mixture. **c)** Putative mixture compositions plotted using the fraction of the top (x-axis) and second (y-axis) lineage. Vertical dashed lines indicate the maximum (0.8) and minimum (0.2) top lineage fraction, while the horizontal dashed line indicates the minimum (0.2) second lineage fraction.

**Figure S12 (cont.).** Samples within the central shaded region passed these thresholds. **d)** Distribution of samples binned by number of UShER defining SNPs discriminating the two SARS-CoV-2 lineages in the mixture. **e)** Correlation between the mean allele fraction of discriminating SNPs and the maximum lineage composition for each sample, stratified by the number of discriminating SNPs. In each facet, the Spearman correlation and corresponding p-value are shown above and the y=x line is shown along with a linear model fit in dark red with margin of error in light grey. **f)** The mean allele fraction of SNPs expected in both mixture lineages for each sample stratified by the number of discriminating SNPs. **g)** Violin plots showing the depth of coverage distributions for non-mixture samples compared with the final set of mixtures.

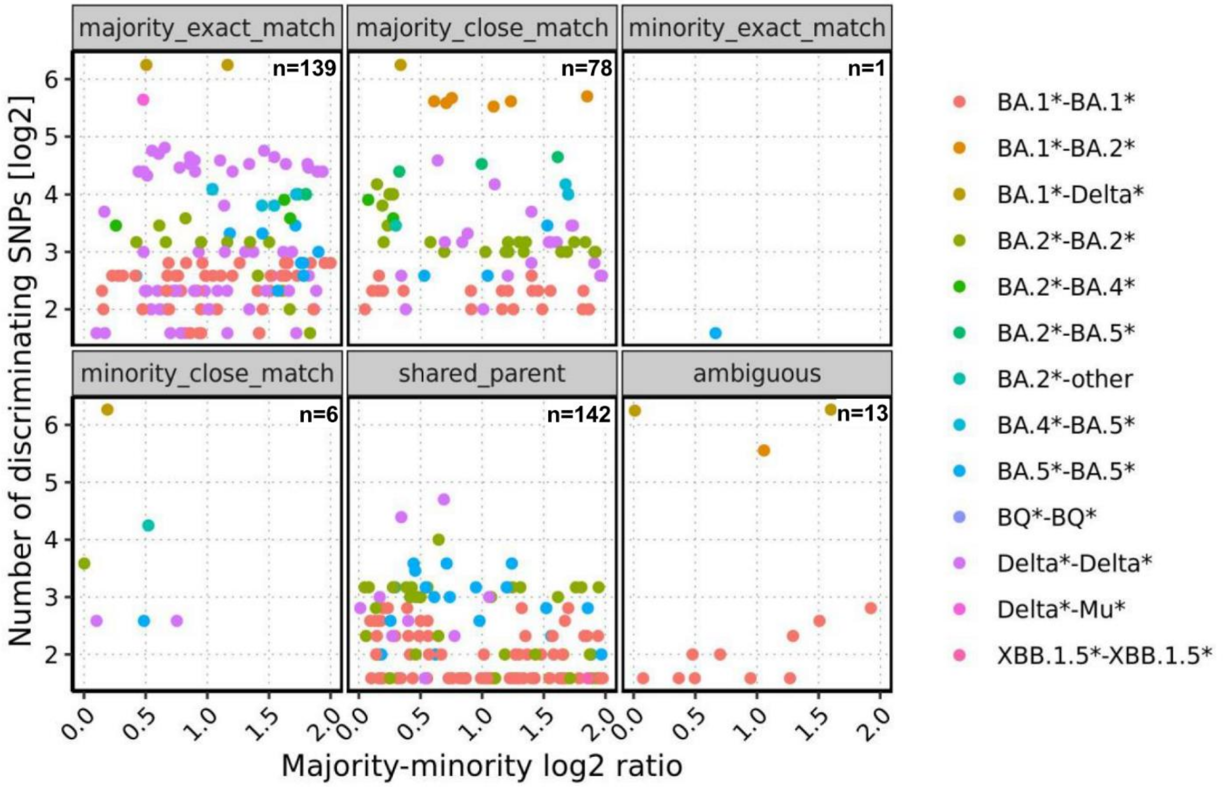


**Figure S13. Concordance between PANGO lineage calls determined by pangolin and the majority/minority mixture lineages determined by freyja.** Each data point represents one of the 379 mixtures colored based on the two lineages comprising the mixture, e.g. a BA.1*-BA.1* mixture contains two distinct BA.1 sublineages. Each mixture is plotted based on the log2 ratio of majority and minority lineage fractions (x-axis) and the number of discriminating UShER defining SNPs (y-axis, log2) and stratified by lineage concordance category. The pangolin/freyja lineage comparisons are categorized as exact if the sublineage determination is precisely the same, whereas close matches indicate that the pangolin lineage is a parent/descendant of one of the freyja lineages. The comparison is labelled as "shared_parent" if the pangolin lineage call is a parent of both the majority and the minority freyja lineage. If the pangolin lineage call is neither a parent nor a descendant of either freyja lineage, then the result is "ambiguous". The total number of mixture samples in each category is shown at the top right of each panel.

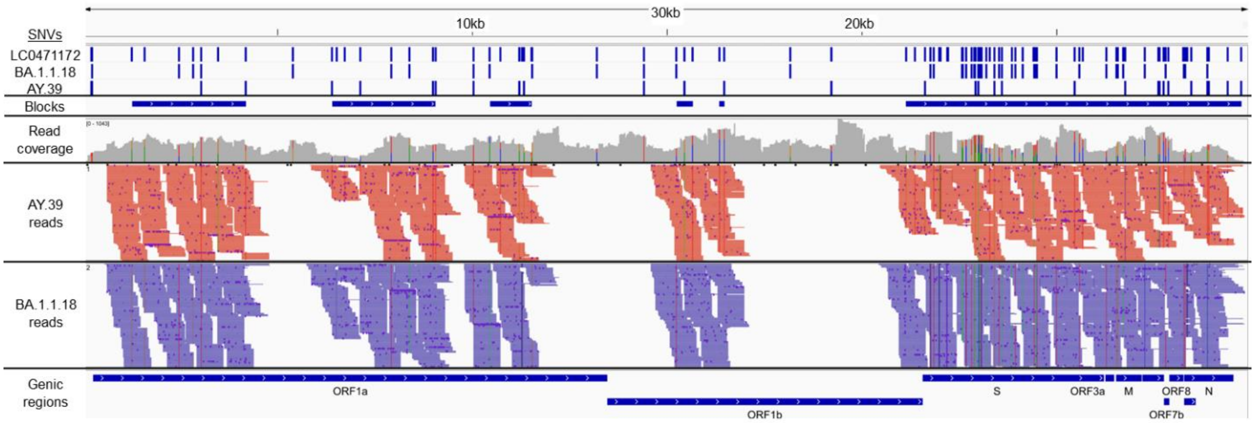


**Figure S14. Integrative Genomics Viewer depiction of sample LC0471172, which had the largest merged haplotype block size (~15.8 kbp).** The CLC mutation calls are shown at the top with the mixture lineage defining SNVs (BA.1.1.18, AY.39) and the original whatshap haplotype blocks below. Total read coverage is shown with the reads assigned to each of the mixture lineages below (red = AY.39, blue = BA.1.1.18). SARS-CoV-2 genic regions and genomic positions are at the bottom and top of the image, respectively.

**Table S1.** Raw and log10-transformed sample counts for each U.S. state and region (District of Columbia).

| State / region | Number of samples | Number of samples (log10) |
|---|---|---|
| Alabama | 2,650 | 3.42 |
| Alaska | 1,958 | 3.29 |
| Arizona | 17,476 | 4.24 |
| Arkansas | 2,838 | 3.45 |
| California | 62,371 | 4.30 |
| Colorado | 6,419 | 3.81 |
| Connecticut | 14,061 | 4.15 |
| Delaware | 5,960 | 3.78 |
| District of Columbia | 8,434 | 3.93 |
| Florida | 36,138 | 4.30 |
| Georgia | 9,817 | 3.99 |
| Hawaii | 1,706 | 3.23 |
| Idaho | 4,578 | 3.66 |
| Illinois | 23,273 | 4.30 |
| Indiana | 11,841 | 4.07 |
| Iowa | 2,899 | 3.46 |
| Kansas | 5,761 | 3.76 |
| Kentucky | 12,980 | 4.11 |
| Louisiana | 1,603 | 3.20 |
| Maine | 291 | 3.00 |
| Maryland | 17,246 | 4.24 |
| Massachusetts | 9,175 | 3.96 |
| Michigan | 4,603 | 3.66 |
| Minnesota | 3,644 | 3.56 |
| Mississippi | 1,667 | 3.22 |
| Missouri | 8,462 | 3.93 |
| Montana | 1,412 | 3.15 |
| Nebraska | 2,295 | 3.36 |
| Nevada | 11,959 | 4.08 |
| New Hampshire | 4,805 | 3.68 |
| New Jersey | 48,182 | 4.30 |
| New Mexico | 10,570 | 4.02 |
| New York | 37,272 | 4.30 |
| North Carolina | 45,777 | 4.30 |
| North Dakota | 67 | 3.00 |
| Ohio | 10,548 | 4.02 |
| Oklahoma | 2,191 | 3.34 |
| Oregon | 9,957 | 4.00 |
| Pennsylvania | 20,889 | 4.30 |
| Rhode Island | 8,154 | 3.91 |
| South Carolina | 8,944 | 3.95 |
| South Dakota | 319 | 3.00 |
| Tennessee | 4,464 | 3.65 |
| Texas | 14,082 | 4.15 |
| Utah | 4,897 | 3.69 |
| Vermont | 239 | 3.00 |
| Virginia | 27,201 | 4.30 |
| Washington | 19,820 | 4.30 |
| West Virginia | 11,417 | 4.06 |
| Wisconsin | 11,033 | 4.04 |
| Wyoming | 209 | 3.00 |
| **Total** | **594,554** | **5.77** |

**Table S2. SARS-CoV-2 mixture lineage group co-occurrence frequencies.** Each entry in the table represents the frequency of two lineage groups co-occurring in a sample, where the most and second-most abundant lineage groups are shown on the left and top, respectively.

| Most abundant lineage group | Second-most abundant lineage group | | | | | | | | | Total |
|---|---|---|---|---|---|---|---|---|---|---|
| | BA.1 | BA.2 | BA.4 | BA.5 | BQ | Mu | Delta | other | XBB.1.5 | |
| BA.1 | 150 | 3 | 0 | 0 | 0 | 0 | 3 | 0 | 0 | 156 |
| BA.2 | 4 | 70 | 2 | 2 | 0 | 0 | 0 | 1 | 0 | 79 |
| BA.4 | 0 | 4 | 0 | 4 | 0 | 0 | 0 | 0 | 0 | 8 |
| BA.5 | 0 | 2 | 2 | 33 | 0 | 0 | 0 | 0 | 0 | 37 |
| BQ | 0 | 0 | 0 | 0 | 1 | 0 | 0 | 0 | 0 | 1 |
| Delta | 3 | 0 | 0 | 0 | 0 | 0 | 92 | 0 | 0 | 95 |
| Mu | 0 | 0 | 0 | 0 | 0 | 0 | 1 | 0 | 0 | 1 |
| other | 0 | 1 | 0 | 0 | 0 | 0 | 0 | 0 | 0 | 1 |
| XBB.1.5 | 0 | 0 | 0 | 0 | 0 | 0 | 0 | 0 | 1 | 1 |
| Total | 157 | 80 | 4 | 39 | 1 | 0 | 96 | 1 | 1 | 379 |
